# UKB-KG: Knowledge Graph for Integrating and Enhancing Biomedical Insights from the UK Biobank

**DOI:** 10.64898/2026.09.02.26361902

**Authors:** Zhe Wang, Jie Wang, Yue Shen, Shan Gao, Yeqiu Chen, Hanzhu Chen, Chuandong Cheng, Kaixian Yu, Hongtu Zhu, Jieping Ye

## Abstract

The UK Biobank (UKB) is a cornerstone of modern biomedical research, providing unparalleled data to advance the understanding, prediction, and treatment of diseases. Its contributions span genetics, genomics, disease prediction, and long-term follow-up studies, driving transformative advancements in public health and precision medicine. However, the fragmentation of research outcomes across numerous publications limits analytic efficiency and cross-study integration. To address this, we developed UKB Knowledge Graph (UKB-KG), a high-quality medical knowledge graph constructed using large language models (LLMs) with 88.8% precision as assessed by GPT-5.4. Integrating data from approximately 9,200 UKB-related publications. UKB-KG comprises 292,328 triples enriched with contextual attributes such as source information and demographic details. It reveals intricate relationships among diseases, genes, chemicals, lifestyle factors, and other biomedical entities, while a dynamic scoring mechanism enhances triple retrieval accuracy. Evaluations highlight UKB-KG’s transformative potential. (i) Embedding UKB-KG into multi-disease prediction models improves AUROC, AUPRC, and F1 scores by 8.1%, 6.2%, and 5.6%, respectively, for rare diseases. (ii) A tailored retrieval-augmented generation (RAG) approach boosted LLM accuracy by 13.2% on PubMedQA. and (iii) A user-friendly platform enhances accessibility for researchers. By unifying fragmented research and enabling robust data exploration, UKB-KG emerges as a powerful tool for advancing biomedical research and driving innovative healthcare applications.

## 1 Introduction

The UK Biobank (UKB) is a leading health research database with longitudinal data on 502,467 UK participants aged 40 to 69, representing diverse ethnic backgrounds, predominantly European, supporting extensive population-based studies (Sudlow et al. 2015; Miller et al. 2016; Bycroft et al. 2018). Its extensive dataset includes phenotypic, genomic, medical, lifestyle, and environmental data, making it invaluable for studying disease progression, identifying risk factors, and advancing personalized medicine, disease prediction, prevention, and large-scale genetic studies. UKB’s longitudinal design allows researchers to track health outcomes and disease trajectories over time, offering critical insights into the interplay of genetics, environment, and lifestyle. However, the dispersion of research outputs across numerous publications poses challenges for efficiently synthesizing and integrating findings (Figure 1). Consolidating this scattered information is essential to fully unlock UKB’s potential, driving deeper insights into disease mechanisms and transformative advancements in medical research and healthcare innovation.

**Figure 1:**
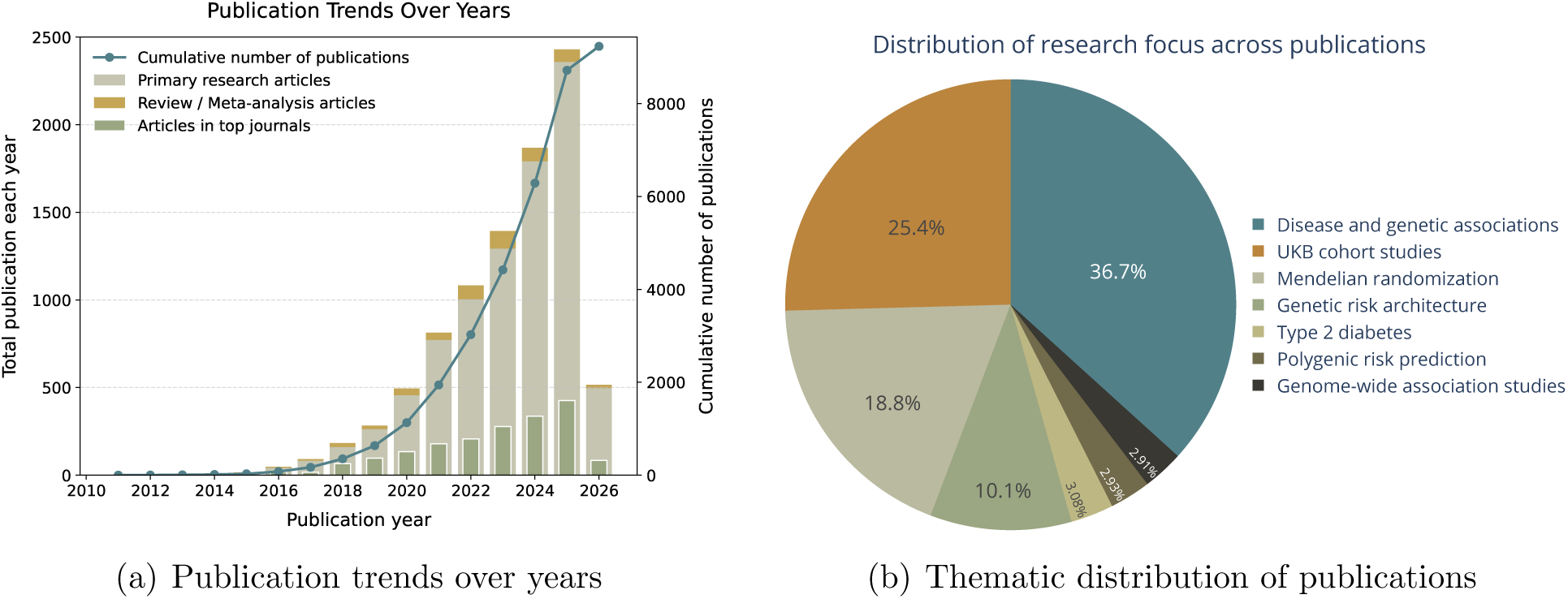
Temporal and thematic insights into highly relevant UKB publications. (a) The publication trends over years (up to March 2026). Annual publication counts are categorized into primary research articles, review/meta-analysis articles, and articles published in top journals (with an impact factor greater than 10). (b) The thematic distribution, categorized into seven major clusters based on their titles and abstracts.

Medical knowledge graphs (MKGs) have become a powerful tool for organizing and utilizing vast, heterogeneous biomedical data (Li et al. 2022; Chandak et al. 2023). By transforming unstructured literature, such as free text and tables, into structured knowledge units: triples, consisting of entities and their relationship. MKGs create a cohesive and navigable network of knowledge. For instance, a study on diabetes and cardiovascular disease may yield a triple: [*diabetes, positively correlates with, cardiovascular diseases*], which helps researchers to quickly understand essential findings in a publication and enables further understanding of connections across publications. Initially, MKG construction relied heavily on manual curation and expert annotations, exemplified by resources like the Unified Medical Language System (UMLS) (Bodenreider 2004), which standardized medical terminologies. While valuable, these approaches were time- and labor-intensive, restricting their ability to scale with biomedical knowledge growing. The integration of natural language processing (NLP) and machine learning (ML) marked a pivotal shift, enabling partial automation and improving scalability through systems like the Biomedical Informatics Ontology System (BIOS) (Yu et al. 2022). More recently, large language models (LLMs) are revolutionizing MKG construction, significantly enhanced the extraction and organization of medical knowledge from vast literature. LLMs enable up-to-date, expansive MKGs, offering unprecedented potential for integrating biomedical knowledge and advancing research and applications.

Existing MKGs face significant challenges. A major limitation is the omission of contextual features that specify the conditions under which triples are extracted, such as source information and baseline characteristics. Source information, including author details, publication date, research institution, and journal, is essential for assessing the credibility and relevance of extracted relationships. Without proper attribution, researchers cannot verify the validity of the knowledge, especially as medical conclusions evolve over time. Baseline characteristics like gender, age, ethnicity, and lifestyle also play a critical role, influencing disease susceptibility, symptoms, and treatment outcomes. For instance, the triple [*severe chest pain, is a precursor to, heart attack*] may primarily apply to men, while women often exhibit atypical symptoms like fatigue. The absence of such demographic context limits MKGs’ relevance for personalized medicine and population-specific research. Furthermore, many automated MKG construction methods fail to fully utilize the semantic capabilities of LLMs. Hallucination effects in LLMs often lead to the extraction of inaccurate or redundant information, compromising precision and reliability. Addressing these contextual gaps and refining LLM integration are critical steps toward improving the accuracy and practical utility of MKGs in biomedical research and healthcare.

This paper introduces **UKB-KG**, a specialized and reliable MKG designed to support diverse UKB-related biomedical studies. Utilizing LLMs, UKB-KG integrates data from around 9,200 high-quality UKB-related publications, including free text and tables, and generates 292,328 relationships among 83,793 entities of 10 types. Especially extended the scope from abstract to highly relevant sections of full text, e.g. results, discussion, tables and etc, for better quantifying the contexts where the triples sit. The UKB-KG construction pipeline (Figure 2, leverages optimization steps such as Triple Filtering and Triple Verification, Entity Alignment and KG Fusion to enable a fully automated, rapid KG construction process with triple extraction precision of 88.8% evaluated by GPT-5.4 (Achiam et al. 2023) and a well-structured topology. Triples are enriched with article-level contextual features derived from their source publications, including 23 attributes for source traceability and 9 baseline characteristics, ensuring relevance to specific populations and enhancing its utility for advanced applications like subgraph analysis and knowledge discovery. A dynamic scoring mechanism further evaluates each triple’s relevance and reliability, ensuring efficient and accurate retrieval.

**Figure 2:**
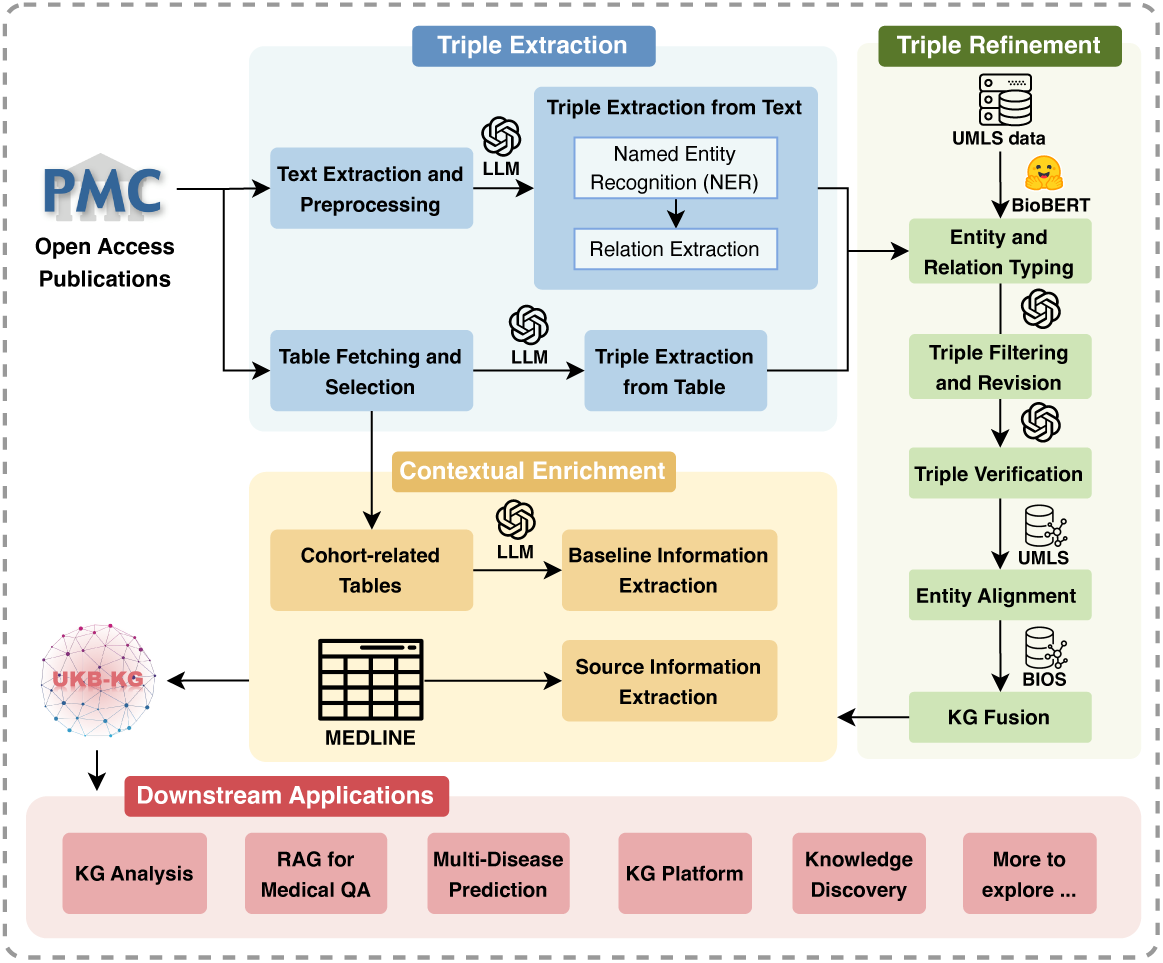
Pipeline of UKB-KG: The entire framework for constructing UKB-KG is divided into three main steps: triple extraction, triple refinement, and contextual enrichment. UKB-KG supports various downstream applications like RAG and multi-disease prediction

We demonstrate UKB-KG’s efficacy through three key applications: **(i) Multi-Disease Prediction:** Integrating UKB-KG embeddings into a prediction model significantly enhances accuracy, achieving a 8.1% improvement in AUROC for rare diseases with limited data. **(ii) Optimized Retrieval-Augmented Generation (RAG):** A graph-based RAG approach tailored for UKB-KG improves PubMedQA benchmark accuracy by 13.2% and 7.8% comparing to baseline LLM and one-shot Chain-of-Thought (CoT), respectively. **(iii) KG Application Platform:** A user-friendly platform providing graph querying and RAG-enhanced chatbot functions, improves accessibility and usability for researchers to the extracted information. By combining LLM-powered automation, contextual enrichment, and innovative applications, UKB-KG established a new standard for MKGs, and enabled more effective utilization of UKB research and driving advancements in biomedical discovery and healthcare innovation.

## 2 UKB-KG: UK Biobank-Knowledge Graph

The large-scale, forward-looking UKB health research project has promoted numerous research projects and published a large volume of high quality discoveries which was chosen as the foundational data source for constructing our knowledge graph (KG), UKB-KG. UKB’s strength lies in the diversity and scale of its data. Its cohort includes over half a million participants, spanning middle-aged to elderly populations, with extensive long-term follow-up data. This unparalleled dataset enables in-depth exploration of complex associations and causal relationships. UKB encompasses over 10,000 variables across genetic data (e.g., whole genome sequencing), imaging data (e.g., MRI of brain, heart, and whole body), lifestyle data (e.g., diet, psychological status, occupation), biological samples (e.g., blood and urine), and health outcomes data (e.g., hospitalizations, cancer diagnoses, mortality). Additionally, UKB covers a wide spectrum of disease states, subclinical conditions, and healthy populations, supporting research across the health-disease continuum. Leveraging this exceptional dataset, thousands of studies have investigated intricate interconnections within biomedical domains. UKB-KG builds on these insights, systematically integrating and structuring relationships to facilitate further applications.

The UKB-KG was constructed end-to-end using GPT-5 (Achiam et al. 2023) with minimal reasoning effort, leveraging 9,187 UKB-related publications (Figure 2 and Section B in the Supplementary Materials). Unlike existing literature-based MKG methods that primarily focus on abstracts, UKB-KG extracted key insights from unstructured texts in abstracts, results, and conclusions (Section B.2.1 - Extraction of Triples from Text), as well as structured tables (Section B.2.2 - Extraction of Triples from Tables). The resulting KG includes 83,793 unique nodes and 292,328 relationships. Entities are categorized into 10 types, based on UKB data and UMLS semantic types (Table 1). Most nodes fall into GENE (21,205), MEAS (measurements; 16,909), and DISO (disease; 14,110), while ACTI (activities/behaviors/phenomena) and BASE (baseline demographics), though smaller, capture essential lifestyle, exposure and population context. A fine-tuned BioBERT classifier, trained on UMLS entities with semantic types reorganized into our predefined entity categories, was employed for entity classification (Section B.3.1 - Triple Typing). As illustrated in Figures 3 and 4, UKB-KG captured complex relationships across data types, primarily centered on DISO.

**Figure 3:**
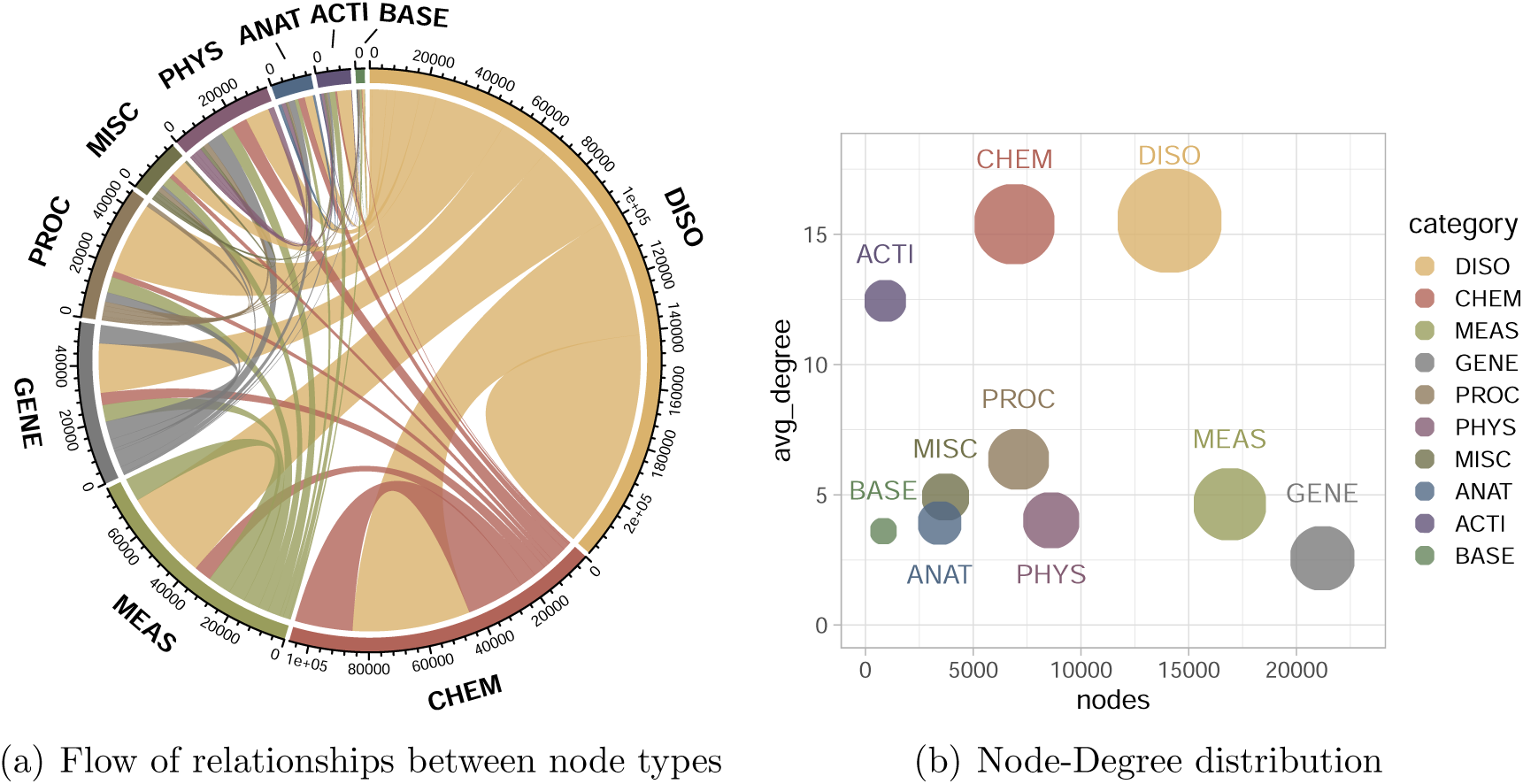
Overview of node types and their relationships in the UKB-KG. (a) The chord plot illustrates the relationships between different types of nodes. (b) The bubble plot shows the distribution of the number of nodes and average degree for each type.

**Figure 4:**
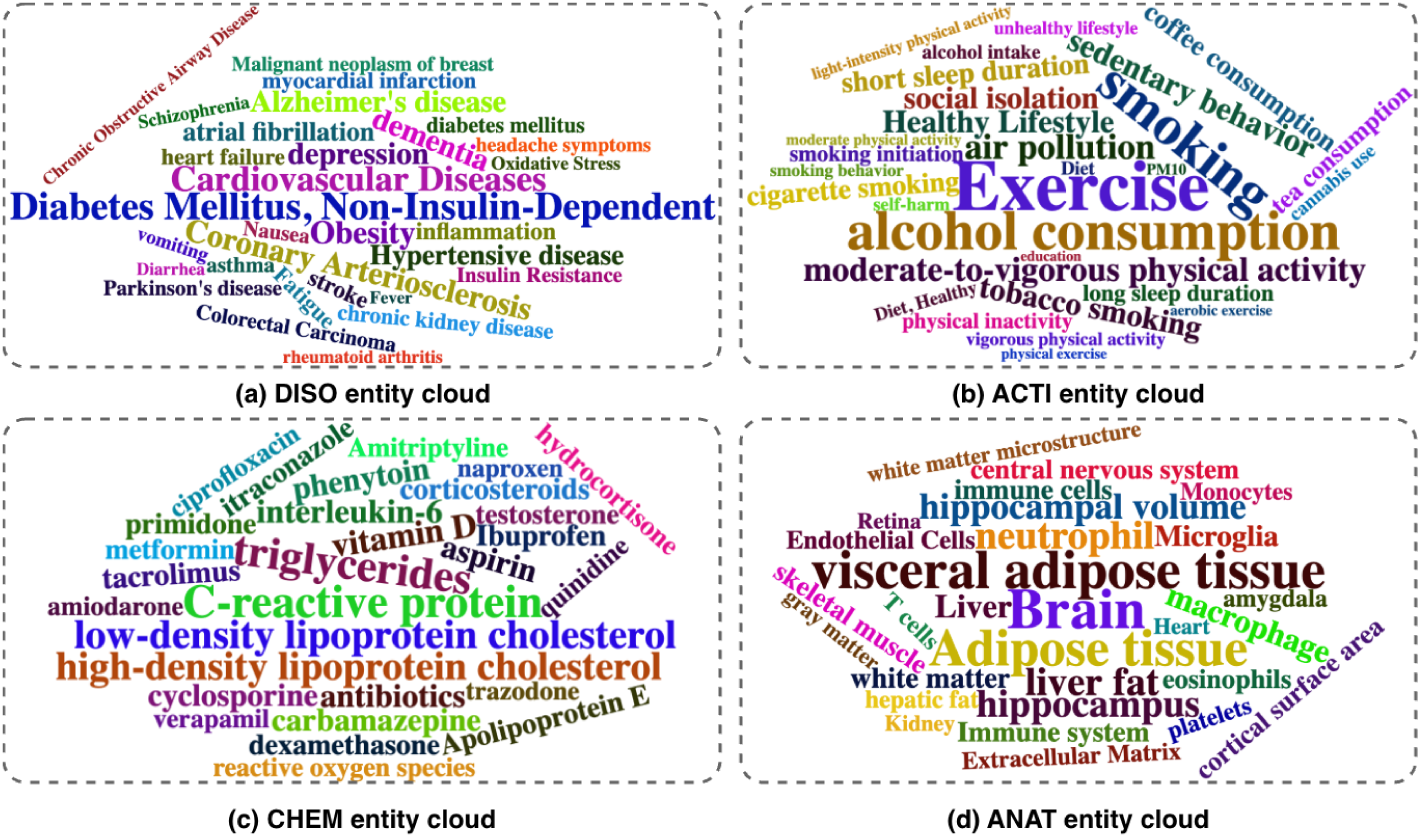
Entity Cloud highlighting key nodes in the UKB-KG across DISO, ACTI, CHEM, and ANAT categories.

**Table 1:** Summary statistics for 10 entity types.

| Abbr | Entity Type | Count | Description |
| --- | --- | --- | --- |
| GENE | Genes & Molecular Sequences | 21,205(25.3%) | Gene or genome; molecular, nucleotide, amino acid, and carbohydrate sequences. |
| MEAS | Measurements | 16,909(20.2%) | Clinical findings, laboratory/test results, and measurable attributes of an organism. |
| DISO | Disorders | 14,110(16.8%) | Disease or syndrome; abnormalities; injury or poisoning; pathologic dysfunction; signs and symptoms. |
| PHYS | Physiology | 8,622(10.3%) | Biologic/physiologic functions and organism/tissue/cell-level functions. |
| PROC | Procedures | 7,106(8.5%) | Procedures in healthcare, diagnosis, laboratory testing, therapy, and research. |
| CHEM | Chemicals & Drugs | 6,918(8.3%) | Chemicals; pharmacologic/biologically active substances; hormones, enzymes, vitamins; hazardous substances; clinical drugs. |
| MISC | Miscellaneous | 3,720(4.4%) | Miscellaneous entities, such as concepts, living beings, objects, devices. |
| ANAT | Anatomy | 3,446(4.1%) | Anatomical structures and regions; body systems/organs; tissues; cells and cell components. |
| ACTI | Activities & Behaviors & Phenomena | 920(1.1%) | Activities, behaviors and phenomena. Many lifestyle and exposure factors are included here. |
| BASE | Baseline Demographics | 837(1.0%) | Baseline demographics, such as population, age, and patient-related groups. |
| Total |  | 83,793 |  |

A distinguishing feature of UKB-KG is that it attaches rich, article-level contextual information to triples, categorized into source information and baseline characteristics (Table 2 and Section B.4 - Contextual Enrichment). These features enhance knowledge traceability and reliability, allowing researchers to evaluate medical conclusions more effectively. This contextual enrichment enables the creation of subgraphs tailored to specific research needs. For example, subgraphs based on publication year can uncover temporal trends in medical findings, facilitating longitudinal analyses. Similarly, demographic-based subgraphs allow exploration of disease traits and associations in specific populations, uncovering insights such as higher disease incidence rates or treatment response disparities.

**Table 2:** Summary of contextual features, including detailed fields and extraction methods.

| Category | Field(Abbr.) | Method |
| --- | --- | --- |
| Source Info | Abstract(AB), Affiliation(AD), Article Identifier(AID), Author Identifier(AUID), Full Author(FAU), Date of Publication(DP), Grant Number(GR), ISSN(IS), Issue(IP), Journal Title Abbreviation(TA), Journal Title(JT), Language(LA), MeSH Terms(MH), Publication Type(PT), PubMed Central Identifier(PMC), PubMed Unique Identifier(PMID), Title(TI), Volume(VI) | MEDLINE |
|  | Keywords(KW) | XML parsing |
|  | URL(URL) | Generated by DOI |
|  | Journal Impact Factor(JIF) | Retrieved by a Python package |
|  | Citations(CT) | Retrieved via the Europe PMC Articles RESTful API |
|  | If use UKB data(UKB) | Identified from publications linked directly to UKB approved projects |
| Baseline Info | Cohort name, Sample size, Mean age, Age distribution, Gender distribution, Racial distribution, Educational attainment, Employment status, Cohort description | Extracted by LLM from tables |

The UKB-KG is poised to make a transformative impact on biomedical research, offering unique insights and applications that distinguish it from existing MKGs. Firstly, UKB-KG provides reliable new discoveries grounded in its comprehensive and robust data foundation. The massive UKB sample size supports statistically robust analyses, mitigating risks of overfitting and false positives that are common in studies with smaller datasets. Its extensive variable coverage offers a holistic view of health and disease mechanisms, avoiding biases introduced by omitting critical confounding factors to reduce the risk of partial or misleading conclusions. Furthermore, the longitudinal follow-up feature introduces a temporal schema, allowing researchers to track health outcomes over time. This capability is essential for distinguishing causal relationships from mere associations, uncovering true etiological factors. In addition to novel discoveries, UKB-KG serves as a high-quality resource for validating, expanding, and refining other MKGs. It can corroborate overlapping relationships proposed in smaller datasets. For example, if a smaller KG suggests an association between a biomarker and a disease, UKB-KG is able to provide robust evidence to confirm or refute the relationship with a larger and more diverse population. Moreover, integrating UKB-KG with other KGs creates a more comprehensive resource. A KG focused on specific diseases, for instance, can be significantly enriched by incorporating UKB-derived relationships, which span a broader spectrum of variables and diseases. This dual role of discovery and validation makes UKB-KG an indispensable tool for advancing biomedical research.

In summary, UKB-KG, with its robust data foundation and optimized construction process, offers a high-precision, comprehensive representation of medical knowledge. Its ability to capture complex relationships and contextual nuances positions it as an invaluable tool for advancing medical research and driving new discoveries.

## 3 Evaluation and Applications

### 3.1 Graph Evaluation and Ablation Study

To assess the quality of the triples and demonstrate the contribution of key components in our graph construction framework, we conducted a series of ablation experiments.

#### Triple Precision Evaluation

We first explored variations in prompt engineering, specifically the number of in-context examples used in the triple extraction prompt (0-shot, 1-shot, and 5-shot), to validate the robustness of the extraction process. In addition, we evaluated the effectiveness of the Triple Verifier (Section B.3.3 - Triple Verification), which is designed to mitigate LLM hallucinations and improve precision. To quantify its impact, we compared extractions performed with and without the Triple Verifier, and further compared against a cross-model verification setting (Shami et al. 2025; Consoli et al. 2025). In the cross-model setting, triple validity is determined by majority voting across multiple models: GPT-5 (Achiam et al. 2023) with minimal effort, and DeepSeek-V4-Flash (DeepSeek-AI 2026) and Qwen3.6-Flash (Qwen Team 2026) in thinking mode. A triple is accepted as correct only when at least two models judge it as correct. We used the following evaluation metrics:

#### Micro Precision

The ratio of correct triples to the total number of extracted triples across the entire dataset, representing the overall precision of the extraction process.

#### Macro Precision

The mean precision of triples extracted from each individual article, which reflects the balanced performance across various articles.

#### Number of Recalls

Computing the conventional recall score is infeasible, as it would require complete human annotation of all articles to obtain ground-truth triples. Such an assumption contradicts the purpose of our work, which is to extract triples through computational methods. Following SAC-KG (Chen et al. 2024), we therefore report the average number of correctly verified triples per article as a denominator-free recall proxy. This type of proxy is commonly used in open-ended information extraction, where exhaustively annotating all valid facts is impractical, as also discussed in Vo and Bagheri 2017 and Kolluru et al. 2020. It serves as a meaningful surrogate for actual recall, reflecting how effectively a method converts the input corpus into validated knowledge.

Building on methodologies established in SAC-KG (Chen et al. 2024), Vicuna (Chiang et al. 2023), and G-Eval (Liu et al. 2023), we employed GPT-5.4 with medium reasoning effort as an automated assessor to evaluate the correctness of triples extracted from text (evaluation prompt: Figure S11). We randomly sampled 500 articles, and summarize the results in Table 3. Overall, the differences among 0-shot, 1-shot, and 5-shot prompting are minor, indicating that extraction quality is largely insensitive to the number of in-context examples, with 1-shot prompting achieving the best overall performance. Removing the Triple Verifier leads to a clear drop in precision, highlighting its role in maintaining relatively high precision. Compared with cross-model majority voting, the Triple Verifier achieves better overall performance, indicating that a single-model verifier can already help mitigate hallucinations to some extent in our framework. The observed reduction in recall is attributed to a decrease in both incorrectly verified triples and redundant triples, which is expected behavior.

**Table 3:** Ablation Study: Evaluation results of triple extraction.

| Model | Micro Precision | Macro Precision | Number of Recalls |
| --- | --- | --- | --- |
| **UKB-KG** (w/ 1-shot) | 88.75 | 88.04 | 15.34 |
| UKB-KG w/ 0-shot | 88.75 | 86.54 | 15.18 |
| UKB-KG w/ 5-shot | 88.18 | 86.95 | 14.17 |
| UKB-KG w/o Verifier | 86.15 | 85.23 | 16.16 |
| UKB-KG w/ CM Verifier | 86.72 | 85.09 | 14.66 |

*w/* and *w/o* denote *with* and *without*, respectively. *CM* denotes *cross-model* verification (majority voting across multiple models).

#### Graph Structure Evaluation

Named Entity Recognition (NER) refines the graph by identifying concise, medically meaningful entities prior to extraction, aiming to reduce inappropriate entities and mitigate cases where the same concept is extracted under multiple surface forms (see Supplementary Section B.2.1). For instance, without NER, the triple [*smoking*, *associated with*, *lower risk of lung cancer*] was extracted. While semantically correct, this triple embeds the relationship within the entities rather than expressing it through the relation, which also generates more derivative entities of “lung cancer”. To evaluate the impact of NER, we conducted a structural analysis on the graphs constructed from text-extracted triples derived from the same 500 sampled articles. As shown in Table 4, enabling NER produces a more compact graph (fewer nodes) while increasing the clustering coefficient and the normalized betweenness centrality, indicating improved cohesion. Figure S2 further corroborates this effect: the post-NER graph exhibits denser connectivity and integrates previously isolated triples into the main network.

**Table 4:** Ablation Study: Evaluation results of graph structure.

| Metric | UKB-KG | UKB-KG w/o NER |
| --- | --- | --- |
| Node Count | 6,221 | 6,491 |
| Average clustering coefficient | 0.0220 | 0.0053 |
| Normalized betweenness centrality (max) | 0.0129 | 0.0010 |
| Normalized betweenness centrality (mean) | $4.44 \times 10^{-5}$ | $1.59 \times 10^{-6}$ |

### 3.2 Multi-Disease Prediction

Multi-disease prediction, which leverages a patient’s medical history to forecast the likelihood of future diseases, is a key area of research in the UKB community. Traditional approaches often simplify medical history variables into binary indicators and use them as input features in machine learning models for multi-label classifications. While effective in some cases, these binary representations result in sparse features and limited training data, particularly for rare diseases. This sparsity restricts the model’s ability to capture complex patterns and associations in the data.

UKB-KG can address these challenges by embedding rich medical knowledge derived from UKB cohort data into predictive models. Knowledge graph embeddings (KGE), characterized by their dense and informative nature, mitigate feature sparsity and reduce the reliance on extensive training samples. By integrating domain knowledge through KGE, models can better capture intricate relationships within the data for rare diseases, significantly improving the accuracy and reliability of multi-disease prediction.

#### Data Description

We curated EHR data for 498,452 participants from the UKB cohort, focusing on medical history variables derived from primary care records, hospital inpatient data, and self-reports. Medical histories were annotated using Phecodes, which map tens of thousands of International Classification of Diseases (ICD) diagnosis codes into higher-level categories (Denny et al. 2010; Wu et al. 2019), streamlining the definition and analysis of disease phenotypes. Timestamps were also recorded for temporal modeling. The objective of this task is to predict the occurrence of diseases in the next six months, based on prior medical histories. For each instance, two time points, *t*_1_ and *t*_2_, were selected as the two most recent medical records with an interval of less than six months. Phecodes recorded on or before *t*_1_ were used as input features, while those recorded at *t*_2_ served as ground truth labels for prediction. After filtering, the final dataset comprised 278,985 instances (the long-tailed distribution of Phecode labels shown in Figure S3), which were randomly divided into training, validation, and test sets in a 7:2:1 ratio.

#### UKB-KGE Enhanced Multi-Disease Prediction

To integrate domain knowledge from UKB-KG into disease prediction, we incorporate KGE into our predictive model by replacing sparse binary inputs with dense graph-based representations. Specifically, each Phecode was mapped to its corresponding node in UKB-KG, and we obtained node embeddings using the KGE model. To simplify the KGE implementation, we mapped all relationships to 12 relations selected from UMLS Semantic Network. For a patient instance, we aggregated the embeddings of all historical Phecodes recorded at or before *t*_1_ by taking their average, yielding a fixed-dimensional representation of the medical history. This KGE-based input captures semantic and relational information from UKB-KG, enriching disease representations beyond co-occurrence signals in the raw EHR.

We evaluate two experimental setups:

- **Baseline**: Uses binary indicators of medical history as input features, which is a 1560-dimensional vector representing the presence (1) or absence (0) of different Phecodes. The output is a 1560-dimensional multi-hot vector representing the predicted diseases.
- **KGE-enhanced**: Uses a dense vector formed by averaging the KGE representations of historical Phecodes as the input, while the output remains the same. To assess robustness to the embedding choice, we evaluate multiple KGE models, including ComplEx (Trouillon et al. 2016), HAKE (Zhang et al. 2020), ModE (Zhang et al. 2020), RotatE(Sun et al. 2019), and TransE (Bordes et al. 2013), with node embedding dimensionality of 1000.

#### Results

We categorized the Phecode labels into four groups based on the number of positive training samples to reflect disease rarity, and evaluated models using macro-averaged AUROC, AUPRC, and F1-score across these groups. To assess statistical significance, we performed paired analyses across Phecode labels using one-sided Wilcoxon signed-rank tests, with bootstrap 95% confidence intervals for the mean paired differences and Benjamini–Hochberg correction for multiple comparisons. As shown in Figure 5 and Table 5, incorporating UKB-KG embeddings consistently improved performance over the binary baseline across all metrics and rarity groups. Importantly, these gains were observed for all evaluated KGE models, indicating that the improvement is primarily driven by the structured knowledge encoded in UKB-KG rather than being sensitive to a specific embedding method.

**Figure 5:**
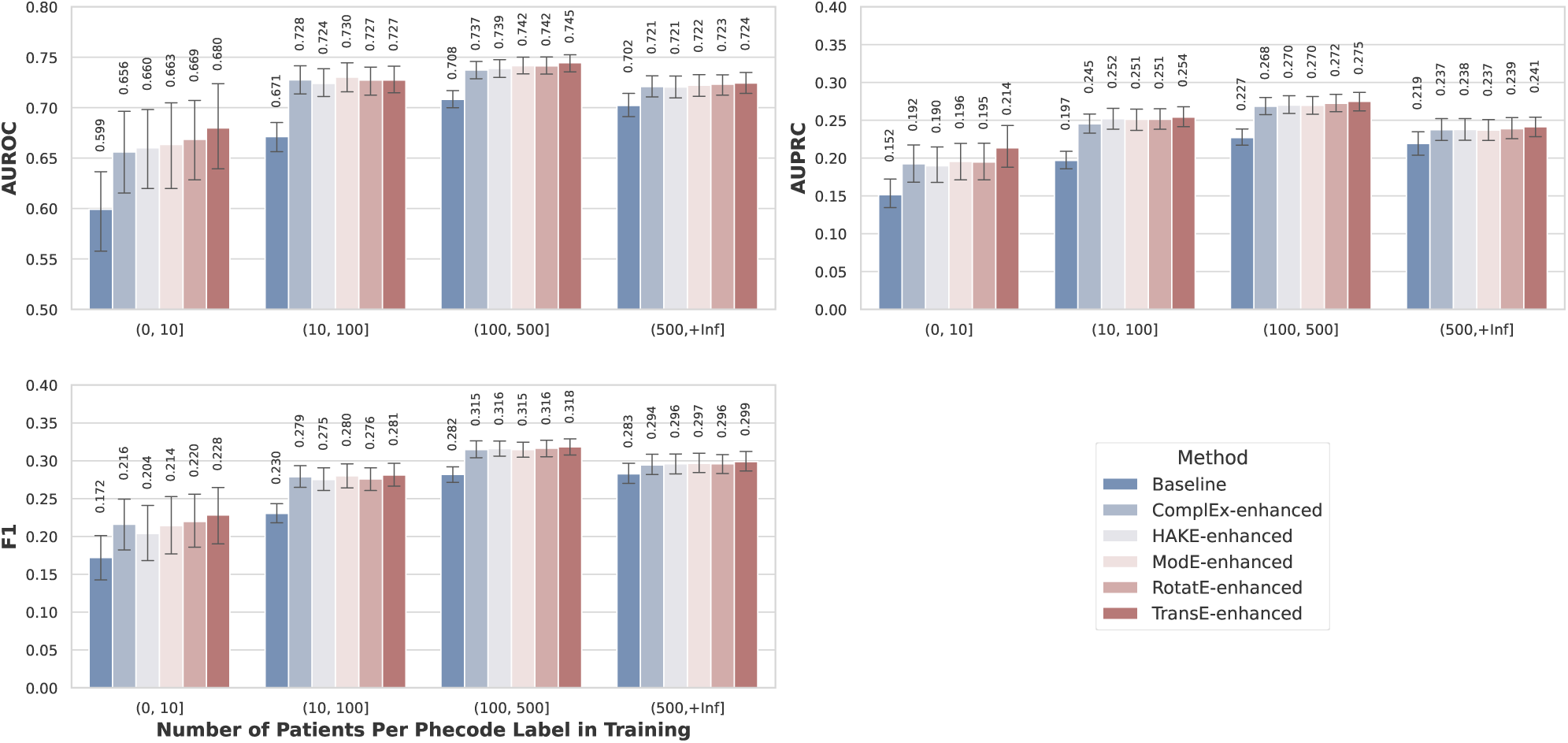
Performance comparison for multi-disease prediction between the binary-history baseline and the model using the average KGE of medical history. Bars show the mean AU-ROC, AUPRC, and F1-score across four phenotype groups stratified by the number of training patients per Phecode label; error bars indicate 95% confidence intervals (ci=95). The UKB-KGE-enhanced models show competitive performance across all phenotype groups, with particularly notable improvements for rare phenotypes.

**Table 5:** Label-level paired comparison of TransE with the binary-history baseline. Δ is the mean paired difference with bootstrap 95% CI; *q* denotes BH-adjusted one-sided Wilcoxon *p*-values.

| Metric | Group | $\Delta$ (95% CI) | $q$ |
| --- | --- | --- | --- |
| AUROC | (0, 10] | +0.081 (0.037, 0.124) | $2.3 \times 10^{-4}$ |
| | (10, 100] | +0.056 (0.043, 0.069) | $< 10^{-10}$ |
| | (100, 500] | +0.036 (0.032, 0.041) | $< 10^{-10}$ |
| | (500, +Inf] | +0.022 (0.019, 0.026) | $< 10^{-10}$ |
| AUPRC | (0, 10] | +0.062 (0.040, 0.085) | $1.5 \times 10^{-6}$ |
| | (10, 100] | +0.057 (0.047, 0.067) | $< 10^{-10}$ |
| | (100, 500] | +0.048 (0.042, 0.054) | $< 10^{-10}$ |
| | (500, +Inf] | +0.022 (0.018, 0.026) | $< 10^{-10}$ |
| F1 | (0, 10] | +0.056 (0.022, 0.091) | $4.4 \times 10^{-4}$ |
| | (10, 100] | +0.051 (0.036, 0.066) | $< 10^{-10}$ |
| | (100, 500] | +0.036 (0.029, 0.044) | $< 10^{-10}$ |
| | (500, +Inf] | +0.016 (0.012, 0.020) | $< 10^{-10}$ |

The improvements were most pronounced in low-sample groups (0,10] and (10,100], where sparse binary histories provide limited supervision. In the most challenging (0,10] group, the KGE-enhanced setting improves AUROC, AUPRC, and F1 by 8.1%, 6.2%, and 5.6%, respectively. This pattern is expected because binary medical-history vectors are dominated by zeros and largely capture surface-level co-occurrence, which becomes noisy and unstable when positives are extremely scarce. In contrast, UKB-KG embeddings provide dense priors that encode semantic and relational context in the graph, enabling better generalization to rare diseases with limited data.

To further contextualize the gains, we also examined performance improvements stratified by intrinsic Phecode system categories. Using TransE as a representative instantiation, the category-wise analysis reveals broadly positive shifts across most categories (Figure 6), indicating that the benefit is not confined to a small subset of disease systems.

**Figure 6:**
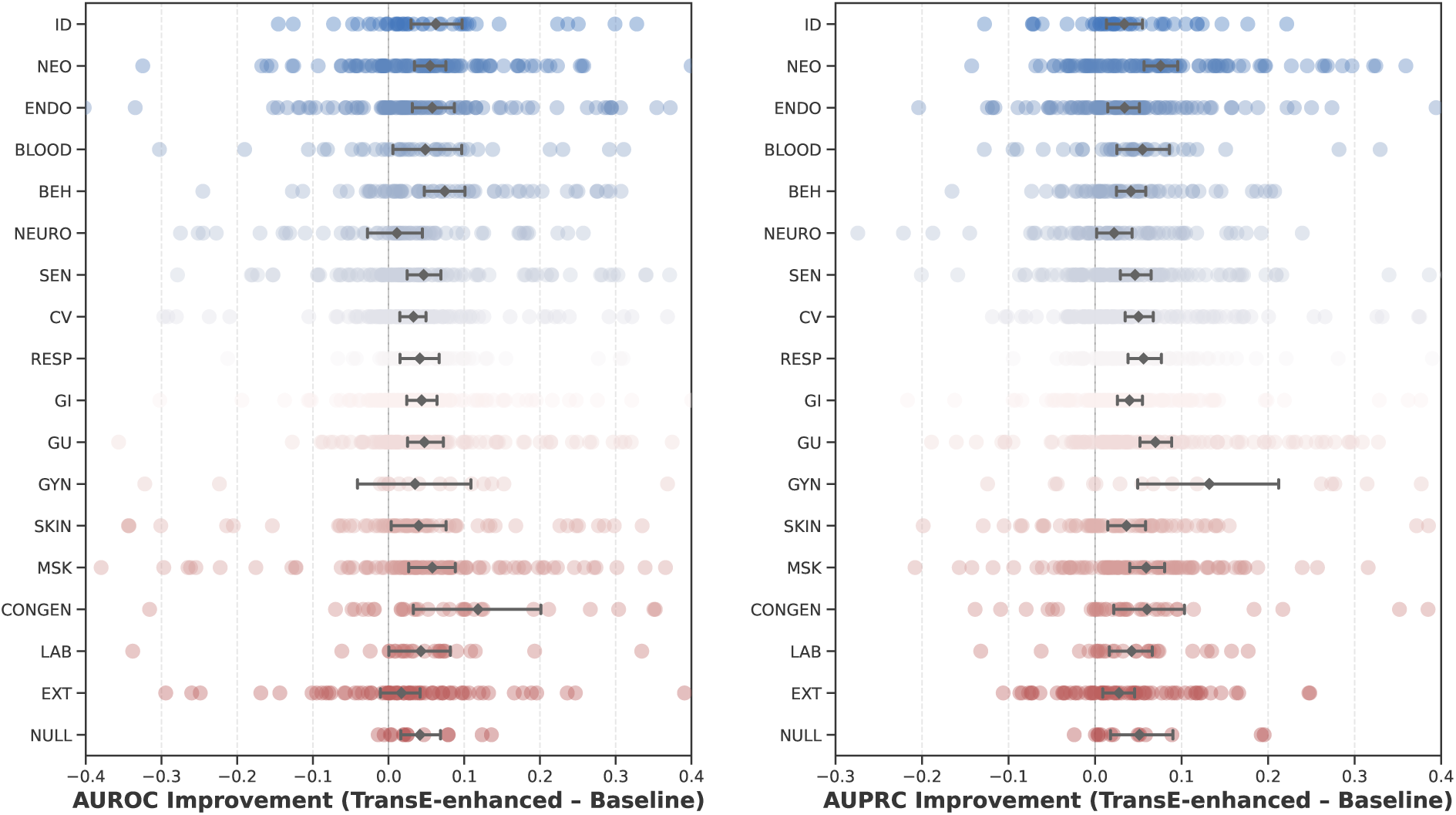
AUROC (left) and AUPRC (right) improvements of the UKB-KGE-enhanced model over the binary baseline across Phecode categories; error bars show 95% confidence intervals. (ci=95).

Overall, the results demonstrate that UKB-KG provides prior knowledge that significantly enhances multi-disease prediction, with benefits that are robust across different KGE instantiations and particularly salient for long-tailed disease labels.

### 3.3 Retrieval-Augmented Generation (RAG)

In medical question answering, LLMs struggle with lack of up-to-date information and specialized medical knowledge; thus, often present outdated and/or inaccurate information. RAG mitigates these issues by integrating real-time retrieval of relevant information from external knowledge sources to enhance response accuracy and reliability. However, retrieving relevant content from vast volumes of information remains a formidable challenge. To harness the potential of UKB-KG, we propose an optimized RAG approach specifically tailored to this KG. Our method combines neighbor-based and path-based retrieval strategies while incorporating triple confidence scores, ensuring that the retrieved information is both relevant and reliable.

The **optimized UKB-KG-based RAG** framework integrates a structured retrieval process followed by a generation process to deliver accurate and relevant medical answers.

Given an input question *q*, we first extract a set of medical entities *E* = *{e*_1_*, e*_2_*, …, e_n_}*. These entities are aligned to UMLS concepts via the method described in Section B.3.4, resulting in *E^′^* = *{e^′^_1_, e^′^_2_, …, e^′^_n_}*. For each aligned entity *e^′^_i_*, the *k*_1_ closest nodes *N_i_* = *{n^i^_1_, …, n^i^k*_1_*}* are retrieved from UKB-KG. Candidate nodes are ranked using cosine similarity between BioBERT-generated embedding vectors.

To ensure comprehensive and relevant retrieval, the process utilizes both a **path-based retriever** and a **neighbor-based retriever**. The **path-based retriever**, designed for questions involving relationships between entities (e.g., “What is the relationship between COVID-19 and diabetes?”), selects the top-*k*_2_ relevant paths from the set of shortest paths (up to 4 hops) between nodes in *N* = *S_n_ N_i_*. The **neighbor-based retriever**, best suited for questions focused on a specific entity (e.g., “What are the effects of COVID-19 on the human body?”), extracts all first-order neighbors of a node *n^i^*, generating a set of candidate triples *T* = *{t*_1_*, t*_2_*, …, t_m_}*.

One major challenge is managing data redundancy due to the large number of retrieved triples. To address this, we introduce a combined scoring mechanism to rank and refine the retrieved triples, calculated as:

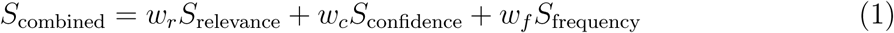

where *S*_relevance_ measures the relevance between the triple and the question using normalized cosine similarity of their BioBERT embeddings, *S*_confidence_ assesses the reliability of the triple, factoring in contextual features such as publication date, citation count, journal impact factor, and the use of UKB data, and *S*_frequency_ reflects the frequency of the triple’s occurrence. The weights *w_r_, w_c_,* and *w_f_* satisfy *w_r_* + *w_c_* + *w_f_* = 1. Details on score calculations are provided in Supplementary Section C.3 - Calculations of scoring for RAG. The retrieved knowledge incorporated as additional context along with the input question is then fed into the LLM to generate the final answer.

#### Datasets

- **PubMedQA**: We use PubMedQA (Jin et al. 2019) as the primary benchmark for our experiments. It targets challenging biomedical research questions and is widely used to evaluate medical knowledge retrieval and reasoning capabilities. Built from PubMed articles, PubMedQA provides questions (article titles), contexts (abstracts without conclusions), long answers (conclusions) and final answers (“Yes,” “No,” or “Maybe”). To evaluate the effectiveness of the RAG tailored for UKB-KG, we provide the question only, without using the original context included in the dataset. Experiments are conducted on the PQA-Labeled test set, which consists of 500 instances.
- **BioASQ**: BioASQ (Krithara et al. 2023) is a biomedical question answering benchmark built from expert-curated questions and PubMed literature. In our experiments, we use the Yes/No questions from the Task B test sets of the most recent three years (2023–2025), yielding a total of 270 questions. We further exclude the dataset-provided relevant articles, assessing performance without curated evidence.

#### Baseline Methods

We employ GPT-5 as the backbone LLM and evaluate our proposed RAG method against the following baseline approaches:

- **GPT-5**: The LLM directly generates answers without any special prompt engineering.
- **Zero-shot/Few-shot CoT**: Chain-of-Thought (CoT) prompting (Kojima et al. 2022; Wei et al. 2022) improves the reasoning ability of LLMs by encouraging them to generate explicit intermediate reasoning steps before arriving at a final answer. In our experiments, we adopt both zero-shot CoT and one-shot CoT as baselines.

#### Results

We evaluate all methods using two metrics: accuracy and macro-F1. As shown in Table 6, our proposed RAG framework consistently outperforms baseline methods on both PubMedQA and BioASQ. On PubMedQA, our method delivers clear improvements over GPT-5 (13.2%) and CoT (9.0% in zero-shot and 7.8% in one-shot). While in BioASQ, zero-shot RAG achieves the best overall results, reaching an accuracy of 0.874 and a macro-F1 of 0.870. These results highlight the effectiveness of UKB-KG in integrating information from the literature and supporting more accurate biomedical question answering.

**Table 6:** Results of RAG on PubMedQA and BioASQ datasets. Baselines include basic GPT-5 and Chain-of-Thought (CoT).

| Dataset | Metrics | GPT-5 | CoT |  | RAG |  |
| --- | --- | --- | --- | --- | --- | --- |
|  |  |  | 0-shot | 1-shot | 0-shot | 1-shot |
| PubMedQA | Accuracy | 0.370 | 0.412 | 0.424 | 0.480 | **0.502** |
|  | Macro-F1 | 0.329 | 0.371 | 0.366 | 0.399 | **0.416** |
| BioASQ | Accuracy | 0.852 | 0.859 | 0.863 | **0.874** | 0.870 |
|  | Macro-F1 | 0.846 | 0.855 | 0.858 | **0.870** | 0.865 |

### 3.4 Interactive Platform for UKB-KG: Querying and Chatting

To enhance accessibility for researchers and general users, we have developed a user-friendly graph platform for viewing and analyzing UKB-KG data. The platform supports complex graph queries, contextual feature exploration, and a RAG Chatbot, enabling efficient access to essential information and insights, even for users with limited technical expertise.

#### Graph Overview

This section provides a high-level overview of UKB-KG, including its purpose, key statistics, and construction methodology, offering users a comprehensive understanding of the graph at a glance.

#### Basic Graph Search

This feature facilitates quick and straightforward queries: (i) *Search Subgraph by Node Name:* Users can retrieve a subgraph of a node’s first-order neighbors in both graphical and tabular formats. For example, Figure S4 shows the query result for “diabetes.” If the node is not found, the platform suggests similar alternatives. (ii) *Search Contextual Features by PMCID:* Users can view all contextual features of a specified article.

#### Advanced Graph Search

The platform supports customizable queries, allowing users to define parameters such as entity names and types, relation properties, and the number of hops. For instance, Figure S5 demonstrates a query that retrieves a subgraph containing up to 100 DISO-type entities linked to *smoking* within 2 hops, with triple properties (e.g., scores) available via interactive display.

#### RAG Chatbot

The RAG Chatbot is designed to enable intuitive engagement with UKB-KG for augmented Q&A. Tailored for individuals without a medical background, it provides additional contextual input to the LLM to enhance responses (Section 3.3 - Retrieval-Augmented Generation (RAG)). Users can explore detailed information supporting the LLM-generated answers, categorized as follows: (i) *Neighbor-based Context:* Includes relevant triples, their PMCIDs, hyperlinks, and source texts, offering a broad perspective on a single entity. (ii) *Path-based Context:* Provides an in-depth exploration of relationships between two entities, visualizing paths for greater clarity. For a detailed demonstration, see Supplementary Section C.4 - Platform Demonstration.

## 4 Conclusion

The UKB-KG framework integrates key insights from diverse UKB literature, producing a refined MKG with 292,328 triples. Optimized LLM processes for extraction, refinement, and contextual enrichment achieve 88.8% GPT-5.4 evaluated accuracy, addressing challenges like data reliability and missing contextual features. The multi-disease prediction model and RAG method built on UKB-KG demonstrate strong performance, while its rich contextual features enable new opportunities in knowledge discovery and personalized healthcare. The graph platform further supports in-depth exploration and analysis, enhancing its utility for biomedical research.

In this work, we deliberately constrained the corpus to UKB-related publications to ensure homogeneity of the underlying evidence base. This design choice offers several advantages: (i) it reduces noise arising from population heterogeneity and cross-study processing differences, (ii) it provides a more focused analytical target centered on a single, deeply phenotyped cohort, and (iii) it preserves the potential to validate novel discoveries against the rich longitudinal data available within UKB itself. However, we acknowledge that this scope limitation may restrict the diversity of captured knowledge and could impact generalizability in downstream analyses. Importantly, the proposed construction framework is designed to be easily extensible. Expanding the corpus to incorporate broader biomedical resources, such as the full PubMed or PMC collections, requires minimal architectural adjustment, enabling users to scale coverage as needed. Furthermore, the core workflow can be readily adapted to other fields such as finance, education, and agriculture, offering a practical pathway toward continuously updated, domain-specific knowledge graphs. Additional discussion of future directions is provided in Supplementary Section D.

UKB-KG, enriched with source information and demographic characteristics, have enhanced knowledge retrieval and traceability. While their potential for understanding disease distributions, diagnostics, and personalized treatments remains underexplored, further investigation could unlock significant benefits for the biomedical research community.

## Data Availability

The data used in this study comprise both publicly available and restricted-access resources. Publicly available resources include the PMC Open Access corpus, PubMedQA, and BioASQ. UMLS data are available upon registration and acceptance of a license agreement. UK Biobank cohort data are accessible only through an application process and are subject to data use agreements. The BIOS knowledge graph is subject to licensing restrictions and cannot be redistributed. Accordingly, the full set of data cannot be made publicly available.

## Acknowledgment

This research has been conducted using the UK Biobank Resource under Application Number 22783, subject to a data transfer agreement. We thank the participants in the UKB study for their contribution and the research teams for the work in collecting, processing, and disseminating these datasets for analysis. We thank the University of North Carolina at Chapel Hill and the Research computing groups for providing computational resources and support that have contributed to the research results. This work was supported in part by National Key R&D Program of China under contract 2022ZD0119801; National Nature Science Foundations of China grants U23A20388 and 62021001; The Joint Fund for Medical Artificial Intelligence under Grant MAI2022Q011.

## 5 Author Contributions

Contributions of co-authors are described following the CRediT, Contributor Roles Taxonomy https://credit.niso.org/. Conceptualization: ZW, JW, HZ, and JY. Methdology: ZW, YS and HC. Data curation: ZW, YS, YC, SG. Downstream tasks: ZW, SG (SG was responsible for the UK Biobank cohort data processing). Resources: JW and HZ. Supervison: JW, CC, KY, HZ, and JY. Writing - original draft: ZW, YS, and YC. Writing - review & editing: all authors.

## References

Achiam, J., Adler, S., Agarwal, S., Ahmad, L., Akkaya, I., Aleman, F. L., Almeida, D., Altenschmidt, J., Altman, S., Anadkat, S., et al. (2023). Gpt-4 technical report. arXiv preprint arXiv:2303.08774.

Bodenreider, O. (2004). The unified medical language system (umls): integrating biomedical terminology. Nucleic Acids Research, 32(suppl 1):D267–D270.

Bordes, A., Usunier, N., Garcia-Duran, A., Weston, J., and Yakhnenko, O. (2013). Translating embeddings for modeling multi-relational data. Advances in neural information processing systems, 26.

Bycroft, C., Freeman, C., Petkova, D., Band, G., Elliott, L. T., Sharp, K., Motyer, A., Vukcevic, D., Delaneau, O., O’Connell, J., et al. (2018). The uk biobank resource with deep phenotyping and genomic data. Nature, 562(7726):203–209.

Chandak, P., Huang, K., and Zitnik, M. (2023). Building a knowledge graph to enable precision medicine. Scientific Data, 10(1):67.

Chen, H., Shen, X., Lv, Q., Wang, J., Ni, X., and Ye, J. (2024). Sac-kg: Exploiting large language models as skilled automatic constructors for domain knowledge graphs. arXiv preprint arXiv:2410.02811.

Chiang, W.-L., Li, Z., Lin, Z., Sheng, Y., Wu, Z., Zhang, H., Zheng, L., Zhuang, S., Zhuang, Y., Gonzalez, J. E., et al. (2023). Vicuna: An open-source chatbot impressing gpt-4 with 90%* chatgpt quality. See https://vicuna.lmsys.org *(accessed 14 April 2023)*, 2(3):6.

Consoli, S., Coletti, P., Markov, P. V., Orfei, L., Biazzo, I., Schuh, L., Stefanovitch, N., Bertolini, L., Ceresa, M., and Stilianakis, N. I. (2025). An epidemiological knowledge graph extracted from the world health organization’s disease outbreak news. Scientific Data, 12(1):970.

DeepSeek-AI (2026). Deepseek-v4: Towards highly efficient million-token context intelligence.

Denny, J. C., Ritchie, M. D., Basford, M. A., Pulley, J. M., Bastarache, L., Brown-Gentry, K., Wang, D., Masys, D. R., Roden, D. M., and Crawford, D. C. (2010). Phewas: demonstrating the feasibility of a phenome-wide scan to discover gene–disease associations. Bioinformatics, 26(9):1205–1210.

Jin, Q., Dhingra, B., Liu, Z., Cohen, W. W., and Lu, X. (2019). Pubmedqa: A dataset for biomedical research question answering. arXiv preprint arXiv:1909.06146.

Kojima, T., Gu, S. S., Reid, M., Matsuo, Y., and Iwasawa, Y. (2022). Large language models are zero-shot reasoners. Advances in Neural Information Processing Systems, 35:22199–22213.

Kolluru, K., Adlakha, V., Aggarwal, S., Chakrabarti, S., et al. (2020). Openie6: Iterative grid labeling and coordination analysis for open information extraction. arXiv preprint arXiv:2010.03147.

Krithara, A., Nentidis, A., Bougiatiotis, K., and Paliouras, G. (2023). Bioasq-qa: A manually curated corpus for biomedical question answering. Scientific data, 10(1):170.

Li, M. M., Huang, K., and Zitnik, M. (2022). Graph representation learning in biomedicine and healthcare. Nature Biomedical Engineering, 6(12):1353–1369.

Liu, Y., Iter, D., Xu, Y., Wang, S., Xu, R., and Zhu, C. (2023). G-eval: Nlg evaluation using gpt-4 with better human alignment. arXiv preprint arXiv:2303.16634.

Miller, K. L., Alfaro-Almagro, F., Bangerter, N. K., Thomas, D. L., Yacoub, E., Xu, J., Bartsch, A. J., Jbabdi, S., Sotiropoulos, S. N., Andersson, J. L., et al. (2016). Multimodal population brain imaging in the uk biobank prospective epidemiological study. Nature Neuroscience, 19(11):1523–1536.

Qwen Team (2026). Qwen3.6-35B-A3B: Agentic coding power, now open to all.

Shami, F., Marchesin, S., and Silvello, G. (2025). Fact verification in knowledge graphs using llms. In Proceedings of the 48th International ACM SIGIR Conference on Research and Development in Information Retrieval, pages 3985–3989.

Sudlow, C., Gallacher, J., Allen, N., Beral, V., Burton, P., Danesh, J., Downey, P., Elliott, P., Green, J., Landray, M., et al. (2015). Uk biobank: an open access resource for identifying the causes of a wide range of complex diseases of middle and old age. PLoS Medicine, 12(3):e1001779.

Sun, Z., Deng, Z.-H., Nie, J.-Y., and Tang, J. (2019). Rotate: Knowledge graph embedding by relational rotation in complex space. arXiv preprint arXiv:1902.10197.

Trouillon, T., Welbl, J., Riedel, S., Gaussier, É., and Bouchard, G. (2016). Complex embeddings for simple link prediction. In International conference on machine learning, pages 2071–2080. PMLR.

Vo, D.-T. and Bagheri, E. (2017). Open information extraction. Encyclopedia with semantic computing and Robotic intelligence, 1(01):1630003.

Wei, J., Wang, X., Schuurmans, D., Bosma, M., Xia, F., Chi, E., Le, Q. V., Zhou, D., et al. (2022). Chain-of-thought prompting elicits reasoning in large language models. Advances in Neural Information Processing Systems, 35:24824–24837.

Wu, P., Gifford, A., Meng, X., Li, X., Campbell, H., Varley, T., Zhao, J., Carroll, R., Bastarache, L., Denny, J. C., et al. (2019). Mapping icd-10 and icd-10-cm codes to phecodes: workflow development and initial evaluation. JMIR medical informatics, 7(4):e14325.

Yu, S., Yuan, Z., Xia, J., Luo, S., Ying, H., Zeng, S., Ren, J., Yuan, H., Zhao, Z., Lin, Y., et al. (2022). Bios: An algorithmically generated biomedical knowledge graph. arXiv preprint arXiv:2203.09975.

Zhang, Z., Cai, J., Zhang, Y., and Wang, J. (2020). Learning hierarchy-aware knowledge graph embeddings for link prediction. In *Proceedings of the AAAI conference on artificial intelligence*, volume 34, pages 3065–3072.

