## Supplementary Material for "UKB-KG: Knowledge Graph for Integrating and Enhancing Biomedical Insights from the UK Biobank"

#### A Related Work

**Traditional MKG Construction** Early MKG constructions relied on manual curation or rule-based techniques, requiring expert annotation or template design for entity and relationship identification. Notable projects like the UMLS and the Systematized Nomenclature of Medicine - Clinical Terms (SNOMED CT) (SNOMED International 2024) utilized these approaches to integrate various biomedical terminologies and clinical taxonomies respectively, emphasizing structured relationships like synonymy and hierarchical connections. Their development and updates depended heavily on expert validation. Additionally, databases like Online Mendelian Inheritance in Man (OMIM) (Hamosh et al. 2005) focused on the associations between human genes and genetic disorders, involving manual annotation and review by geneticists. Ontology systems like Gene Ontology (GO) (Ashburner et al. 2000) relied on community contributions followed by expert review and approval, to ensure ongoing updates and expansion. While these methods ensured high accuracy and semantic consistency, they were time-consuming, costly, and lacked scalability.

**Automated MKG Construction** Automated methods reduce human labor by applying NLP and ML techniques to various stages of graph construction, including entity and relation extraction, normalization, and graph completion. The semantic knowledge base SemMedDB (Kilicoglu et al. 2012) utilizes the NLP system SemRep (Kilicoglu et al. 2020) to automatically extract semantic predications (subject–predicate–object) from PubMed literature. Rotmensch et al. (Rotmensch et al. 2017) employed three probabilistic models to automatically learn disease-symptom associations from electronic medical records. Beyond basic entity and relation extraction, F. Li et al. (Li et al. 2019) fine-tuned BioBERT for entity normalization using electronic health records (EHR) and several standard datasets. The COVID-19 KG (Zhang et al. 2021) applied state-of-the-art graph embedding methods to complete the graph and identify potential new therapeutic targets or pathological mechanisms related to COVID-19.

Despite these advancements, many KGs still rely on existing ontologies and apply ML only to parts of the construction process. Recently, the BIOS (Yu et al. 2022) introduced a fully algorithmic pipeline, constructing a large-scale biomedical KG without pre-existing ontologies, enabling the generation of new concepts and relationships.

**MKG Construction with LLMs** The advent of LLMs, such as ChatGPT, has opened new possibilities for MKG construction. With their ability to comprehend and generate complex biomedical content with minimal supervision, LLMs are well-suited for constructing MKGs from vast amounts of unstructured text. AutoRD (Cao et al. 2024) is an end-to-end system designed for automatically extracting rare disease information from clinical texts to construct KG by combining LLMs with medical ontologies. ReguloGPT (Wu et al. 2024) leverages GPT-4 to extract molecular regulatory pathways from biomedical literature using in-context learning (ICL). DALK (Li et al. 2024) first constructs an Alzheimer's Disease specific KG from relevant literature using LLM, and then retrieves information from the graph to augment the LLM's reasoning capabilities. More recently, MDKG(Gao et al. 2025) proposes an LLM-powered framework for constructing a large-scale, contextualized mental disorders knowledge graph, which incorporates contextual attributes such as conditional statements and demographic factors into triples.

Despite advancements, the modeling and utilization of contextual features in MKG construction remain underexplored. Furthermore, to the best of our knowledge, no prior work specifically targets knowledge integration and graph construction for UKB research.

### B Methods

#### B.1 Overview

With around 9,200 UKB-related papers, we developed a comprehensive standardized process (shown in Figure 2 in the main text) to extract information from UKB literature, involving three parts: **(i) Triple Extraction** This part focus on extracting triples from the key information in the heterogeneous data of literature including texts and tables. Our approach harnesses the capabilities of LLMs to understand and extract specialized knowledge from biomedical texts based on In-Context Learning (ICL) including instruction, few-shot examples and response format. Moreover, these steps also make use of NLP tools like NER to improve the extraction quality. **(ii) Triple Refinement:** This part refines the extracted triples through five steps. Entity typing assigns semantic categories to entity mentions, improving the consistency and interpretability of the extracted triples. Triple filtering and revision remove low-quality triples and revise non-canonical expressions to improve overall triple quality. Triple verification employs LLM self-correction to eliminate hallucinated triples. Entity alignment to normalize the entities and KG fusion to enhance the completeness of the KG. **(iii) Contextual Enrichment:** Contextual features are essential for assessing the reliability of extracted triples and characterizing the conditions under which they hold. We focus on two main areas: source information from corresponding papers and baseline characteristics of the studied cohort. The former can be retrieved from the MEDLINE (National Library of Medicine (NLM) 2024), while the latter is typically found in the specific tables.

#### B.2 Triple Extraction

##### B.2.1 Extraction of Triples from Text

The detailed steps of triple extraction from text is shown in Figure S1.

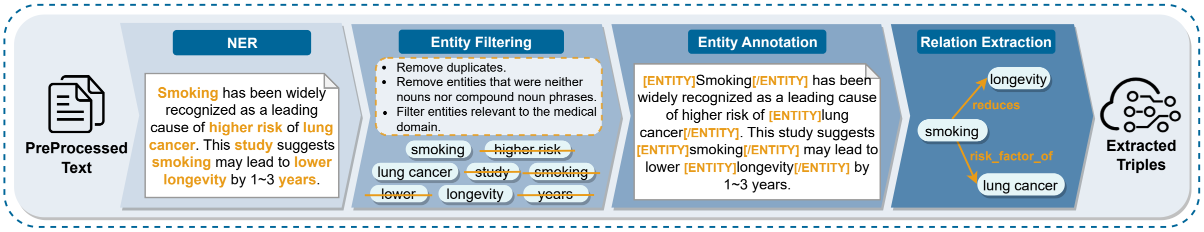


Figure S1: Detailed steps of Triple Extraction.

**Named Entity Recognition (NER)** We first focused on extracting meaningful medical entities while removing duplicates and entities unrelated to medicine. It was observed that LLMs tend to rigidly adhere to the original text, often resulting in extracted triples that match the phrasing of the source verbatim. This can lead to a single entity being fragmented into multiple different forms. For instance, from the sentence "This study suggests smoking leads to higher risk of lung cancer," an LLM might extract the triple [*smoking, lead_to, higher risk of lung cancer*], where "higher" implied a trend. However, it would be more appropriate to reflect such trends in the relationship itself, yielding [*smoking, increases_risk_of, lung cancer*]. A more critical example is the extraction of [*smoking*, *shows*, *strong association with lung cancer*] instead of the more concise and accurate [*smoking*, *associated_with*, *lung cancer*]. Additionally, without further intervention, LLMs also tend to generate overly detailed or meaningless entities.

Therefore, for pre-processed text, we first utilized *en_core_sci_scibert* (Neumann et al. 2019), a specialized NER model trained on scientific and biomedical texts, to extract medical entities. These entities were then refined through a three-step filtering process: (i) removing duplicates, (ii) excluding entities that were neither nouns nor compound noun phrases, and (iii) using the LLM to filter entities relevant to the medical domain. Finally, the filtered entities were annotated within the corpus using `<ent>` and `</ent>` tags to explicitly provided the positional information of the entities.

**Relation Extraction** With the scaling of model size and data size, LLMs show impressive In-Context Learning (ICL) ability, that is, learning from a few examples in the context without additional training. Accordingly, our prompts consist of the task instructions, input data, output format constraints, and optionally a one-shot example.

The final prompt for extracting triples from texts is presented in Figure S8, with the preprocessed text serving as the corpus. The LLM's response yielded a JSON string containing the triples.

##### B.2.2 Extraction of Triples from Tables

Existing literature-based MKGs mainly utilizing free-text paragraphs, especially abstracts, to extract triples, while they often neglected valuable details hidden in structured data like tables. These tables often consolidate key findings—such as risk factors for specific diseases and genetic variations associated with phenotypes—into compact, standardized formats, making them a valuable complementary source of relational knowledge.

To leverage this information, we adopt an LLM-based workflow that treats table understanding as a two-stage task: (i) identify which tables contain extractable biomedical relationships, and (ii) extract triples only from the selected tables. Concretely, we first convert XML tables into readable Markdown text, preserving the caption, body, and footnotes to retain essential context. The LLM then filters out non-informative tables and keeps those expressing meaningful biomedical or genetic relations. Finally, we apply a second LLM pass to extract structured triples from the retained tables, enabling us to incorporate table-derived evidence into UKB-KG while reducing noise from irrelevant tabular content.

#### B.3 Triple Refinement

##### B.3.1 Triple Typing

Biomedical terminology contains many concepts that appear similar on the surface but differ in meaning. Classifying nodes and relationships to suitable types is a crucial step to standardize triple semantics. Moreover, a well-structured KG enhances the efficiency of information querying and retrieval, such as retrieving genetic factors linked to a particular disease.

**Entity Type Schema** UMLS provides fine-grained Semantic Types, and Semantic Groups further aggregate them into coarser categories. Building on this design and considering UKB-specific data characteristics, we predefined 10 entity types in UKB-KG: Genes and Molecular Sequences (GENE), Measurements (MEAS), Disorders (DISO), Physiology (PHYS), Procedures (PROC), Chemicals and Drugs (CHEM), Anatomy (ANAT), Activities/Behaviors/Phenomena (ACTI), Baseline Demographics (BASE), and Miscellaneous (MISC). Among these, MEAS and BASE are introduced as new types to better reflect commonly used UKB variables. Table 1 in the main text summarizes the definitions and statistics of each type. This classification ensures consistent representation and retrieval of medical information. For example, *Microalbuminuria* denotes a clinical measurement (classified as MEAS), whereas *Microalbumin* refers to the substance albumin itself (classified as CHEM).

**BioBERT-based Entity Typing** We trained a BioBERT-based classifier to assign each extracted entity mention to one of the 10 predefined types. Training data were constructed directly from UMLS: to obtain labeled instances for each category, we reorganized UMLS Semantic Types to our 10-type schema and then randomly sampled UMLS concepts and their associated terms according. We split the dataset into training/validation/test sets with a 90:5:5 ratio, partitioned by unique UMLS concepts to avoid leakage (i.e., terms from the same concept do not appear in multiple splits). The resulting dataset contains approximately 4.9 million labeled instances, and the classifier performance on the held-out test set is reported in Table S1.

**Relationship Typing** Relationship types are defined as the pair of the head and tail entity types, represented as `head_type-tail_type` (e.g., `GENE-DISO`).

Table S1: BioBERT entity typing performance on the held-out test set.

| Type | Precision | Recall | F1 | Support |
|---|---|---|---|---|
| ACTI | 0.9592 | 0.9692 | 0.9641 | 3274 |
| ANAT | 0.9835 | 0.9886 | 0.9860 | 3495 |
| BASE | 0.9623 | 0.9634 | 0.9637 | 1429 |
| CHEM | 0.9704 | 0.9488 | 0.9595 | 3772 |
| DISO | 0.9741 | 0.9663 | 0.9702 | 4159 |
| GENE | 0.9701 | 0.9875 | 0.9787 | 6483 |
| MEAS | 0.9879 | 0.9879 | 0.9879 | 6020 |
| MISC | 0.9515 | 0.9348 | 0.9431 | 2563 |
| PHYS | 0.9797 | 0.9773 | 0.9785 | 3958 |
| **Macro avg** | 0.9710 | 0.9695 | 0.9702 | 35153 |
| **Weighted avg** | 0.9734 | 0.9734 | 0.9734 | 35153 |
| **Accuracy** | | 0.9734 | | 35153 |

##### B.3.2 Triple Filtering and Revision

The triples extracted by LLMs often contain substantial noise, including irrelevant content, malformed structures, and non-standard expressions. To improve the overall quality of the extracted triples, we introduce a two-step post-processing procedure consisting of filtering and revision. The prompts used in these two steps are shown in Figures S9 and S10.

**(1) Filtering.** We first remove triples that are either uninformative or poorly formed. This step includes both semantic-based and rule-based filtering:

- **Semantic-based filtering.** We prompt the LLM to identify and discard low-quality triples while retaining only informative biomedical knowledge triples. In particular, this step removes triples that are semantically unclear, poorly defined, low-value, or non-informative, and also deduplicates semantically equivalent triples.

- **Rule-based filtering.** We further apply a set of rules to eliminate badly formatted triples. For example, we remove triples whose entities contain abnormal special symbols or consist entirely of non-alphabetic characters. We also discard triples with entities longer than eight words, as such entities often contain excessive or irrelevant details and are therefore more likely to introduce noise.

**(2) Revision.** After filtering, we further prompt LLM to revise low-quality or non-standard triples to improve their consistency and canonical form, while preserving their factual correctness and original meaning as much as possible. Specifically, we rewrite overly complex relations into more concise and standardized forms, remove redundant descriptive details (e.g., statistical values) from overly specific entities, and expand abbreviations into their full forms when the model can determine them with high confidence.

##### B.3.3 Triple Verification

Althoug LLMs now have developed strong text comprehension capabilities, errors may still occur during KG construction, particularly due to unsupported inferences or knowledge hallucination (Zhang et al. 2023). To improve the factual reliability of the extracted knowledge, we introduce a Triple Verifier based on LLM self-validation.

Specifically, each extracted triple is re-evaluated by the LLM with reference to the original source text. A triple is retained only if it is clearly supported by the text. In this way, the verification step serves as an additional quality control mechanism to reduce hallucinations and improve the factual accuracy of the final KG.

###### B.3.4 Entity Alignment

It is common for terms to exhibit polymorphisms within the context, a phenomenon not unique to biomedical literature but one that poses significant challenges to information retrieval. To better align medical entities in our KG with their formal names, we examined the UMLS, which comprises approximately 3.31 million concepts and 15.7 million concept names derived from 185 distinct source vocabularies. Each concept identifier in UMLS is linked to a canonical name and an extensive array of synonyms, making it a valuable resource for biomedical entity normalization. For UMLS linking, we used ScispaCy tool (Neumann et al. 2019) and applied a similarity threshold of 0.98 to balance precision and recall.

###### B.3.5 KG Fusion

Even after entity normalization, a subset of triples in UKB-KG remains isolated from the main network. To improve connectivity and enrich relational coverage, we augmented UKB-KG with relevant triples from the large-scale BIOS knowledge graph. We first identified candidate UKB-KG entities that can be mapped to BIOS concepts, and then retrieved BIOS relations for candidate entity pairs that lack a corresponding link in UKB-KG, excluding edges *is_a* and *reverse_is_a*. To facilitate retrieval and differentiate the newly incorporated triples from the original ones, we annotate each mapped entity with its BIOS concept identifier (HeadCID/TailCID) and tag each imported relation with the corresponding BIOS relation identifier (RELID).

#### B.4 Contextual Enrichment

In addition to extracting key conclusions as triples from articles, we extracted contextual features corresponding to each source article of a triple, treating these as relational attributes for each triple. Table 2 in the main text depicts that the contextual features are classified into two categories:

- Source Information: detailing the article's origin.
- Baseline Information: describing the study population's demographics and characteristics.

These extra information enabled researchers to quickly assess the relevance, significance, and potential quality of the literature, thereby offering an indirect measure of the derived triples' quality. For instance, the collection of source information aided the creation of academic databases and knowledge bases, facilitating research retrieval. The acquisition of baseline information is crucial for locating literature related to specific target populations, essential for evaluating applicability of extending current research to a wider audience.

##### B.4.1 Source Information Enrichment

To ensure the traceability of the triples for evaluating their accuracy and reliability, we extracted detailed metadata from corresponding articles. The complete list of source information, along with the extraction methods, is provided in Table 2 in the main text. Most information was obtained from the MEDLINE Data Element (Field) (National Library of Medicine (NLM) 2024), which contained specific details about each article to support organization and retrieval. Keywords were extracted by parsing the XML files of the articles.

The URL for each article was constructed by appending the DOI to https://doi.org/. Journal impact factors were retrieved using the *search-impact-factor* Python package based on eISSN, while citation counts were sourced from Scopus (Elsevier 2024). To identify whether an article utilized UKB data, a word-mapping method was employed, classifying an article as using UKB data (mark UKB filed as 1) if "UK Biobank" or its variations appeared in the title or keywords.

Publication date in the source information provides a practical entry point for leveraging the longitudinal nature of UKB, which continuously collects prospective cohort data to support long-term studies. As follow-up duration increases, later studies may refine earlier findings or even report different conclusions; associating triples with publication time therefore enables temporal analyses of how evidence accumulates and evolves. Moreover, although UKB-KG does not directly encode participant-level longitudinal records, it indirectly reflects UKB's temporal advantages through the cohort-derived literature. Many included studies explicitly incorporate temporal dimensions such as follow-up duration, repeated measurements, or cohort progression. Extracting triples from these studies preserves such signals at the level of evidence provenance.

##### B.4.2 Baseline Information Enrichment

Baseline information is typically presented in dedicated tables summarizing cohort demographics and baseline study characteristics. Similar to our table-based triple extraction, we formulate baseline information extraction as a two-stage LLM-based process: (i) identifying tables that present study cohort demographics or baseline characteristics, and (ii) extracting cohort-level demographic information in a structured and standardized format. For tables containing multiple cohorts or subgroups (e.g., cases vs. controls), each cohort is extracted separately together with its corresponding cohort description.

##### B.5 Scalability and Implementation

UKB-KG construction is dominated by API-based LLM calls, and the overall cost scales approximately linearly with the number of processed articles. Table S2 reports the approximate input token usage per valid article for each major LLM-based stage, which can serve as a practical reference for budgeting. Beyond API usage, the BioBERT classifier for entity typing can be fully fine-tuned on a single RTX-3090, while other refinement steps (e.g., entity alignment and KG fusion) are comparatively lightweight.

To accelerate the construction in large-scale deployments, we recommend increasing parallelism at the article level (processing multiple papers concurrently) or batching API requests whenever possible. For self-hosted inference with open-source LLMs, using a high-throughput serving engine such as vLLM can substantially improve parallel decoding efficiency and reduce end-to-end latency.

Table S2: Approximate input token usage per valid article for major LLM-based stages in UKB-KG construction.

| **Stage** | **Tokens per valid article (roughly)** |
|---|---|
| Triple extraction from text | 4.5k |
| Triple extraction from tables | 3k |
| Triple filtering and revision | 1.5k |
| Triple verification (from text) | 2.5k |
| Triple verification (from table) | 2.5k |
| Table selection | 3k |
| Baseline information extraction | 2.5k |

#### C Additional Results

##### C.1 Graph Evaluation and Ablation Study

**Triple Precision Evaluation** As discussed in Section 3.1 in the main text, computing a standard recall score is infeasible because exhaustively annotating all ground-truth triples is impractical in open-ended information extraction settings. We therefore report the average number of correctly verified triples per article (denoted as *Number of Recalls*), which is a commonly used recall surrogate. In addition, we provide an approximate estimate of true recall as a reference. Specifically, we randomly sampled 50 papers and used GPT-5.4 with medium reasoning effort to extract triples from text as a proxy ground truth, which also underwent the procedures of triple filtering and revision, and self-verification. We then computed recall against this proxy ground truth. The estimated recall results are reported in Table S3.

Table S3: Estimated recall using GPT-5.4 as proxy ground truth

| Model | Recall |
|---|---|
| **UKB-KG (w/ 1-shot)** | 61.44 |
| UKB-KG w/ 0-shot | 64.07 |
| UKB-KG w/ 5-shot | 56.10 |
| UKB-KG w/o Verifier | 62.27 |
| UKB-KG w/ CM Verifier | 58.39 |

**Graph Structure Evaluation** Figure S2 visualizes the graphs constructed from the same set of 500 sampled articles with and without the NER component, highlighting its impact on graph connectivity and structure.

#### C.2 Long-tailed Distribution of Diseases

The participants from UKB used in the multi-disease prediction tasks exhibit a long-tailed distribution of Phecode labels (Figure S3).

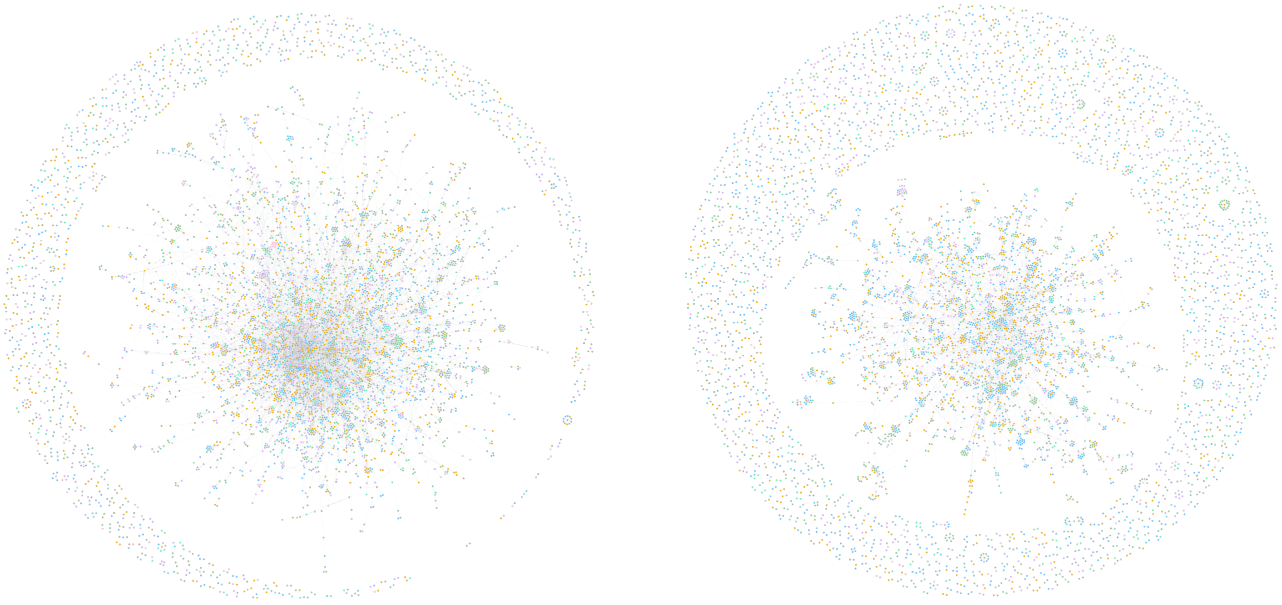

Figure S2: Graph constructed from 500 sampled articles by UKB-KG (left) and UKB-KG without NER (right).

#### C.3 Calculations of scoring for RAG

We introduce a scoring mechanism for the neighbor-based retriever in RAG:

$$S_{\text{combined}} = w_r S_{\text{revelance}} + w_c S_{\text{confidence}} + w_f S_{\text{frequence}} \tag{1}$$

The detailed calculation is outlined below:

1. **Relevance Score** In the RAG framework, the primary criterion for filtering redundant triples is their relevance to the given question, as this directly affects the quality and precision of the generated answer. To achieve this, we employ BioBERT as the embedding model to calculate the cosine similarity between the embedding vectors of each triple and the question. The computed similarity is subsequently normalized to determine the relevance score:

$$S_{\text{relevance}} = (\text{sim}_{\cos}(q, t_i) + 1)/2 \tag{2}$$

2. **Confidence Score** The precision and reliability of a triple are directly proportional

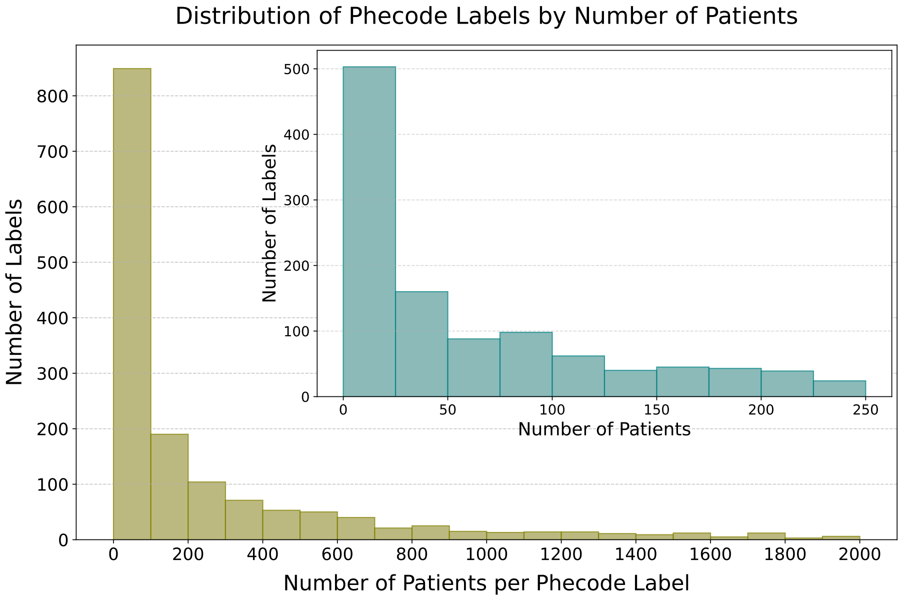


Figure S3: The long-tailed distribution of Phecode labels.

to the confidence in the research articles from which it is derived. To evaluate this, we developed a Confidence Score that employs a holistic set of contextual features to assess the reliability of scientific articles, with particular emphasis on those incorporating data from the UK Biobank. As shown in Equation 3, this metric synthesizes elements including publication date, citation count, journal impact factor, and the utilization of UKB data.

$$S_{\text{confidence}} = w_{dp} S_{\text{DP}} + w_{ct} S_{\text{CT}} + w_{if} S_{\text{IF}} + w_{ukb} S_{\text{UKB}} \tag{3}$$

- **Publication Time Score ($S_{\mathbf{DP}}$)**: Biomedical knowledge evolves rapidly, and newer studies may refine or overturn earlier conclusions. Therefore, more recent evidence is generally more valuable, calculated using the formula

$$S_{\text{DP}}(t) = e^{-\lambda(T-t)} \tag{4}$$

where $\lambda$ is the decay factor, $T$ is the current year, and $t$ is the year of publication.

- **Citation Score ($S_{\mathbf{CT}}$)**: Citations provide a coarse but commonly used proxy for community attention and post-publication scrutiny; highly cited work has typically been examined by a larger audience, which often correlates with higher practical credibility. It is represented by:

$$S_{\text{CT}}(c) = \frac{\log(c+1)}{\log(c_{\max}+1)} \tag{5}$$

where $c$ is the citation count, and $c_{\max}$ is the maximum citation count in the dataset.

- **Impact Factor Score ($S_{\mathbf{IF}}$)**: While imperfect, journal selectivity and editorial standards are frequently correlated with impact factor, which can be used as an auxiliary indicator of publication rigor. It is calculated by:

$$S_{\text{IF}}(i) = 1 - e^{-\alpha i} \tag{6}$$

with $\alpha$ as the decay factor, and $i$ as the journal's impact factor.

- **UKB Score ($S_{\mathbf{UKB}}$)**: Explicit utilization of UKB data is both (a) a signal of methodological strength (large cohort size, rich phenotyping, standardized data collection) and (b) a direct indicator of relevance to the scope of our UKB-centered graph. It is defined as:

$$S_{\text{UKB}} = \begin{cases} 1 & \texttt{if UKB data is used} \\ 0 & \texttt{if UKB data is not used} \end{cases} \tag{7}$$

3. **Frequency Score** The frequency of a triple reflects its prevalence within the knowledge graph. Triples with higher frequency typically indicate that they are more commonly found in medical literature, suggesting stronger generalizability and applicability. The frequency score can be calculated using the following formula:

$$S_{\text{frequency}}(i) = \frac{f_i}{N} \tag{8}$$

Table S4: Evaluations of the combined score weights for our RAG under 0-shot and 1-shot settings. $w_r$, $w_c$, and $w_f$ denote the weights for relevance, confidence, and frequency scores, respectively.

| Dataset | Weights $(w_r, w_c, w_f)$ | 0-shot RAG | | 1-shot RAG | |
|---|---|---|---|---|---|
| | | Accuracy | Macro-F1 | Accuracy | Macro-F1 |
| PubMedQA | (0.90, 0.05, 0.05) | **0.484** | 0.413 | 0.478 | 0.390 |
| | (0.85, 0.10, 0.05) | 0.480 | 0.399 | **0.502** | **0.416** |
| | (0.80, 0.15, 0.05) | 0.480 | **0.416** | 0.476 | 0.384 |
| BioASQ | (0.90, 0.05, 0.05) | 0.867 | 0.863 | 0.867 | 0.862 |
| | (0.85, 0.10, 0.05) | **0.874** | **0.870** | **0.870** | **0.865** |
| | (0.80, 0.15, 0.05) | 0.867 | 0.862 | **0.870** | **0.865** |

where $f_i$ represents the frequency of the $i$-th triple, and $N$ is the maximum frequency among all triples.

In addition to comparisons with baseline methods in the main text, we examine whether incorporating the confidence score and frequency score in the final combined score improves RAG performance. Specifically, we vary the weights of relevance ($w_r$), confidence ($w_c$), and frequency ($w_f$) while keeping the retrieval and generation pipeline unchanged. As shown in Table S4, moderately increasing the weight of confidence is beneficial for better context retrieval quality and improving downstream QA performance.

#### C.4 Platform Demonstration

Figure S4 shows a basic search for "diabetes", while Figure S5 presents the customized settings available for advanced search.

Figure S6 shows the Chatbot's response to "What diseases are related to COVID-19?" accompanied by Neighbor-based Context, which includes *COVID-19*-related triples, associated PMCIDs, hyperlinks, and source texts. Figure S7 presents the answer to "Are diabetes related to smoking?" accompanied by Path-based Context, highlighting potential direct or indirect relationships between *diabetes* and *smoking*, such as their connection through *delta age* and *finding of body mass index*.

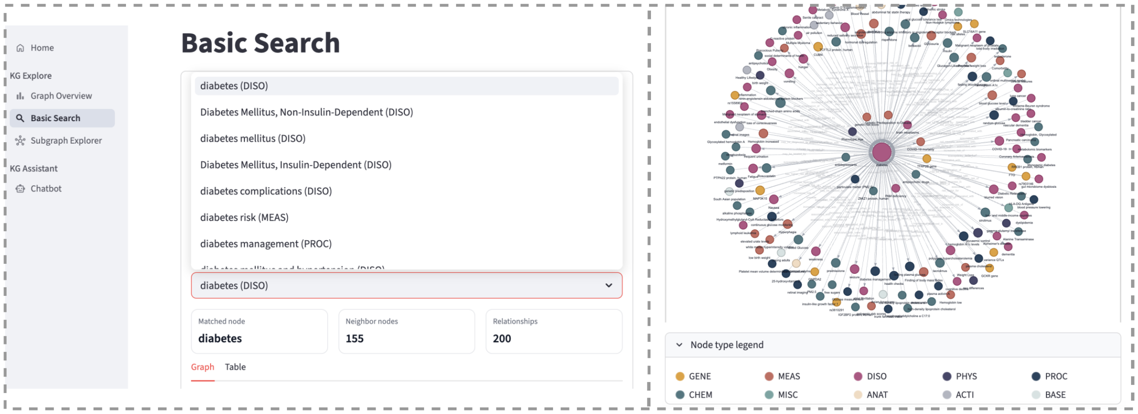


Figure S4: Basic search example

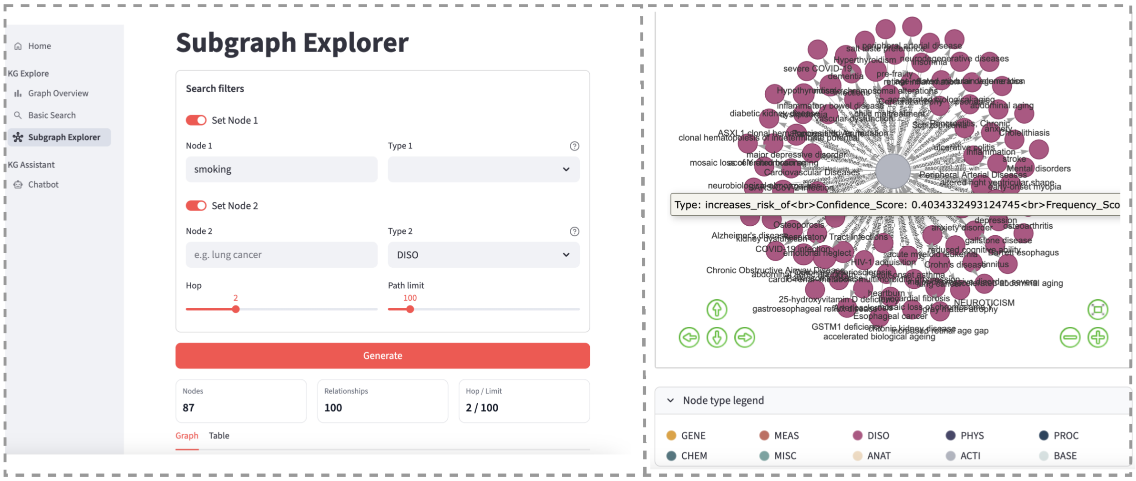


Figure S5: Advanced search example

### D Limitations and Future Work

We summarize the main limitations of UKB-KG and outline directions for future improvement:

**Corpus scope.** We focus on UKB-related publications, motivated by the unique value of UK Biobank and the generally rigorous nature of UKB-based findings. However, this choice

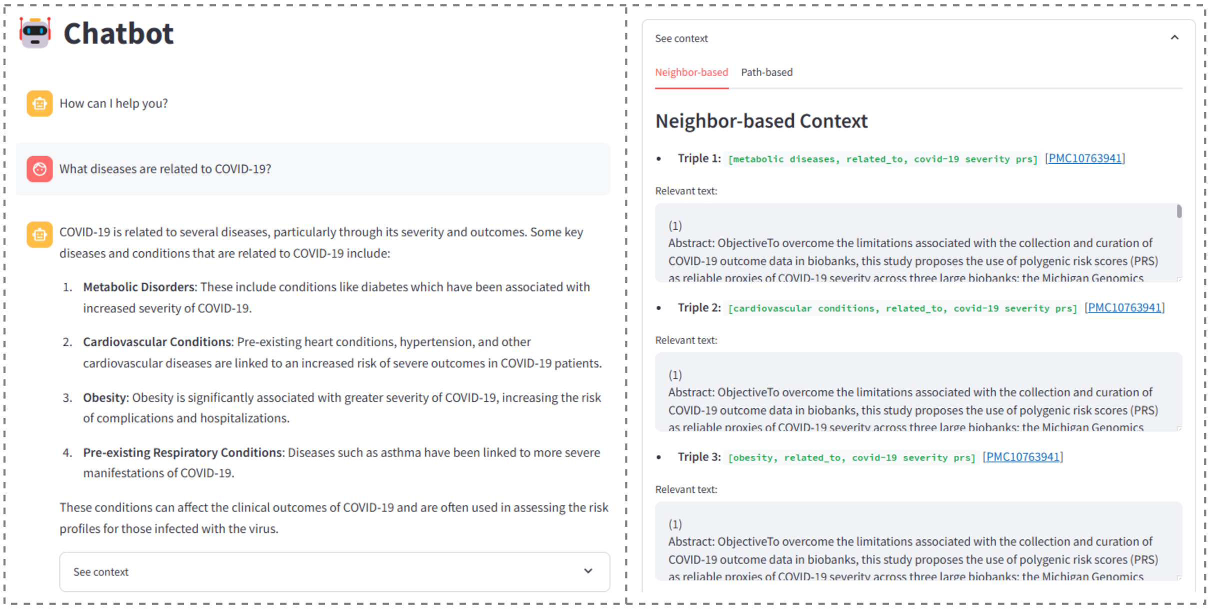


Figure S6: RAG Chatbot example - Neighbot-based Context

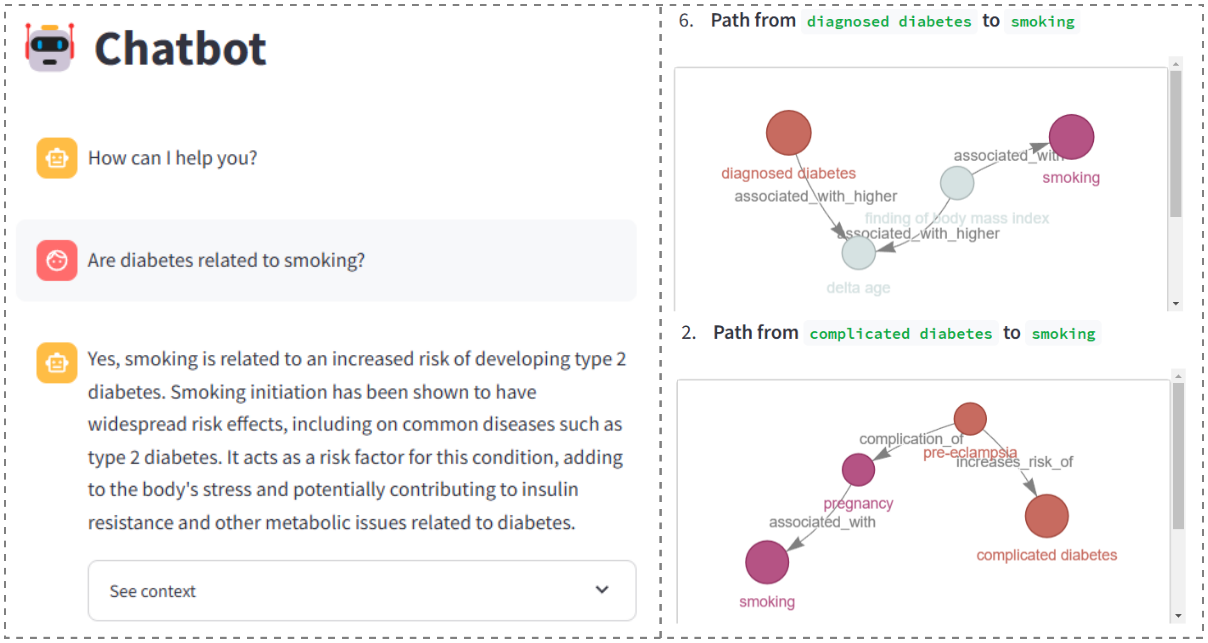


Figure S7: RAG Chatbot example - Path-based Context

inevitably constrains the coverage of entities and relations to what is represented in the UKB-centric literature. Future work could enhance completeness and utility by integrating broader but well-curated external resources (e.g., authoritative domain databases and high-quality biomedical corpora, and carefully filtered general resources when appropriate) together with stronger provenance and harmonization strategies.

**Interpretability beyond provenance.** We have taken steps to improve interpretability and traceability through source information enrichment (explicit provenance), confidence scoring (a quantitative, model-independent estimate of evidential strength), and self-verification (an additional validation pass before inclusion). Nonetheless, interpretability can be further strengthened. We encourage future work to explore: (i) fine-grained evidence anchoring (e.g., linking triples to supporting text spans or table cells), (ii) structured extraction justifications in a constrained format, and (iii) improved uncertainty calibration so that confidence better reflects expected correctness under different relation types and study designs.

**Bias and confounding in observational evidence.** Although UKB is among the most comprehensive biomedical resources available, its observational nature may still introduce biases and confounding factors. To mitigate this concern, future work may consider: (i) incorporating additional large-scale, well-curated datasets (as above) to diversify evidence sources; (ii) evidence typing to distinguish observational associations, predictive modeling results, and causal-inference-oriented analyses (e.g., Mendelian randomization or triangulation when available); (iii) bias auditing tools that surface uneven coverage across demographic groups or phenotypes and flag relations potentially sensitive to population structure or selection effects; and (iv) contradiction and heterogeneity tracking to highlight when later studies refine, qualify, or reverse earlier findings—particularly important for longitudinal, continuously evolving cohorts such as UKB.

### E Prompts

**Triple Extraction from Text**

**# INSTRUCTION**

--------------------

You are a researcher skilled in summarizing scientific findings into concise and informative triples to construct a medical knowledge graph.

The corpus is provided in *TEXT* with medical entities marked between <ent> and </ent>. Your task is to extract meaningful medical triples from *TEXT*.

1. Triple Structure: (Entity1, Relation, Entity2)
 - Entity1 and Entity2 are the medical entities annotated with <ent> and </ent> in the text.
 - Relation represents the semantic relationship between Entity1 and Entity2, inferred from the context.
2. Guidelines for Extraction:
 - Focus on key findings reported in the research. Exclude relationships that do not represent meaningful biomedical knowledge, such as:
   - abbreviation definitions (e.g., [A, abbreviation_of, B]),
   - vague or non-informative relations (e.g., [diabetes, associated_with, diseases], [BMI, is, risk factors], [risk factors, yields, risk]),
   - expressions that do not convey a clear biomedical fact (e.g., [loss, of, noxious heat sensation]).
 - Ensure that the extracted triples reflect factual statements supported by the text, rather than speculative or hypothetical statements.
 - Occasionally, an entity that should remain a single term may be incorrectly split into multiple entities in the text. In such cases, merge them into the correct complete entity when extracting triples. For example, "<ent>breast</ent> <ent>cancer</ent>" should be extracted as "breast cancer".
 - Prefer full names of entities rather than abbreviations whenever possible.
 - If two triples express essentially the same meaning (e.g., due to different relation wording or reversed entity order), extract only one of them.

**# EXAMPLES**

--------------------

- Example text:

````
```
<ent>smoking</ent> has been widely recognized as a leading cause of higher risk of <ent>lung cancer</ent>.... This study suggests <ent>smoking</ent> leads to lower <ent>longevity</ent> by 1~3 year
```
````

- Example response:

````
```json
{
  "Triples": [
    {"Entity1": "smoking", "Relation": "risk_factor_of", "Entity2": "lung cancer"},
    {"Entity1": "smoking", "Relation": "decreases", "Entity2": "longevity"}
  ]
}
```
````

**# TEXT**

--------------------

Here is the text for extraction. Please extract the medical triples and output them in the specified format:

````
```
<<text>>
```
````

Figure S8: The prompt of extracting triples from text.

**Triple Filtering**

**# INSTRUCTION**

--------------------

You are given a set of triples extracted from a scientific article.
Your task is to filter out low-quality triples and keep only valuable biomedical knowledge triples.

**# FILTERING RULES**

--------------------

1. Remove semantically unclear or poorly defined triples.
   Filter out triples whose meaning is unclear, incomplete, or not interpretable as a valid biomedical relation.
   For example: [ CKD | of_uncertain | aetiology ], [ hypertension | highlights_in_models | retinal vessels ]

2. Remove low-value or non-informative triples.
   Discard triples that do NOT convey meaningful biomedical knowledge or research findings, including but not limited to:
   - Trivial or obvious statements
   - Abbreviation or definition relations
   - Statements lacking scientific or biomedical significance
   - Methodological or meta-level statements (not biomedical findings)
   For example: [ age | interrelated_with | risk factors ], [ health data | contributes_to | mental healthcare ], [admixture | affects_reliability_of | methods], [post-trauma stress disorder | is | PTSD], [familial kidney failure | occurs_in | families], [genomic variants | regulate | genes]

3. Deduplicate semantically equivalent triples.
   If multiple triples express essentially the same meaning: keep only ONE representative triple and remove the others.

**# OUTPUT REQUIREMENTS**

--------------------

- Output ONLY the filtered triples
- Include all valid triples that should be retained
- Do NOT modify the wording of triples

**# CANDIDATE TRIPLES**

--------------------

Each triple is structured as: [ head | relation | tail ]

<<triples>>

Figure S9: The prompt of filtering triples.

#### Triple Revision

**# INSTRUCTION**
--------------------
You are given a set of triples extracted from a scientific article.
Your task is to refine low-quality or non-standard triples while preserving their factual accuracy and original meaning as much as possible.

**# REFINE RULES**
--------------------
1. Refine relations:
 If a relation is overly complex, overly specific, overly long, or otherwise non-standard, rewrite it into a more concise and canonical form.

2. Refine entities：
 - Entities must be biomedical entities.
 - If an entity is overly long or overly specific with redundant information, remove redundant descriptive details (such as statistical values) that are not necessary for preserving the correctness of the triple.
 - Prefer full names over abbreviations when you are confident about the expansion and the full name fits naturally in context.

3. Preserve accuracy:
 - Ensure that the refined triple remains factually correct. Do not overstate, strengthen, or exaggerate the original relation or entities beyond the evidence.
 - Only make refinements that improve clarity, normalization, or biomedical correctness.

4. Remove unrecoverable triples:
 - If a triple is too unclear, malformed, or semantically invalid to be reliably refined, remove it.

**# REFINE EXAMPLES:**
--------------------
- [ retinal vessels | used_by_algorithms_to_predict | age ] -> [ retinal vessels | associated_with | age ]
- [ CKD | presents_in_retina_as | sparse capillaries ] -> [ chronic kidney disease | associated_with | retinal capillary rarefaction ]
- [ rs7412 | is_a_variant_in_the_gene | APOE ] -> [ rs7412 | located_in | APOE ]
- [ metformin | leads_to_lower | HbA1c levels ] -> [ metformin | decreases | HbA1c ]
- [ CRP | is_higher_among_patients_with | rheumatoid arthritis ] -> [ C-reactive protein | elevated_in | rheumatoid arthritis ]
- [ APOE4 | increases_the_risk_for_developing | Alzheimer’s disease ] -> [ APOE4 | increases_risk_of | Alzheimer's disease ]

**# OUTPUT REQUIREMENTS**
--------------------
- Keep acceptable triples unchanged. Refine only when necessary.
- For refined triples, ensure the revised version is accurate and standardized.
- Remove triples that are unacceptable and cannot be reliably refined.

**# TRIPLES**
--------------------
Each triple is structured as: [ head | relation | tail ].

<<triples>>

Figure S10: The prompt of triple revision.

```
Triple Evaluation

# INSTRUCTION
--------------------
A corpus from a medical paper is provided in the *INPUT_TEXT*, and the triples extracted from the text are provided in the *EXTRACTED_TRIPLES*. Your task is to evaluate whether each triple is "Correct" or "Incorrect" based on the text.

# EVALUATION_CRITERIA
--------------------
- Correct: The relationship expressed in the triple is clearly supported by the information in the original text.
- Incorrect: The relationship expressed in the triple is not supported by the information in the original text.

# INPUT_TEXT
--------------------
<<text>>

# EXTRACTED_TRIPLES
--------------------
Each triple is structured as: head|relation|tail
<<triples>>

# RESPONSE_FORMAT
--------------------
Evaluate each extracted triple ONE BY ONE based on the *EVALUATION_CRITERIA* and categorize it as "Correct" or "Incorrect."
The evaluation results must follow the exact order of the input triples, and must adhere to the following JSON schema:
```json
{
  "Evaluations": [
    {"Result": "Correct or Incorrect"},
    {"Result": "Correct or Incorrect"},
    ...
  ]
}
```
```

Figure S11: The prompt of evaluating triples.

### References


Ashburner, M., Ball, C. A., Blake, J. A., Botstein, D., Butler, H., Cherry, J. M., Davis, A. P., Dolinski, K., Dwight, S. S., Eppig, J. T., et al. (2000). Gene ontology: tool for the unification of biology. *Nature Genetics*, 25(1):25–29.

Cao, L., Sun, J., and Cross, A. (2024). Autord: An automatic and end-to-end system for rare disease knowledge graph construction based on ontologies-enhanced large language models. *arXiv preprint arXiv:2403.00953*.

Elsevier (2024). Scopus Database. Accessed: 2024-10-28.

Gao, S., Yu, K., Yang, Y., Yu, S., Shi, C., Wang, X., Tang, N., and Zhu, H. (2025). Large language model powered knowledge graph construction for mental health exploration. *Nature Communications*, 16(1):7526.

Hamosh, A., Scott, A. F., Amberger, J. S., Bocchini, C. A., and McKusick, V. A. (2005). Online mendelian inheritance in man (omim), a knowledgebase of human genes and genetic disorders. *Nucleic Acids Research*, 33(suppl_1):D514–D517.

Kilicoglu, H., Rosemblat, G., Fiszman, M., and Shin, D. (2020). Broad-coverage biomedical relation extraction with semrep. *BMC Bioinformatics*, 21:1–28.

Kilicoglu, H., Shin, D., Fiszman, M., Rosemblat, G., and Rindflesch, T. C. (2012). Semmeddb: a pubmed-scale repository of biomedical semantic predications. *Bioinformatics*, 28(23):3158–3160.

Li, D., Yang, S., Tan, Z., Baik, J. Y., Yun, S., Lee, J., Chacko, A., Hou, B., Duong-Tran, D., Ding, Y., et al. (2024). Dalk: Dynamic co-augmentation of llms and kg to answer alzheimer's disease questions with scientific literature. *arXiv preprint arXiv:2405.04819*.

Li, F., Jin, Y., Liu, W., Rawat, B. P. S., Cai, P., Yu, H., et al. (2019). Fine-tuning bidirectional encoder representations from transformers (bert)–based models on large-scale electronic health record notes: an empirical study. *JMIR Medical Informatics*, 7(3):e14830.

National Library of Medicine (NLM) (2024). MEDLINE/PubMed Data Element (Field) Descriptions. Accessed: 2024-10-28.

Neumann, M., King, D., Beltagy, I., and Ammar, W. (2019). ScispaCy: Fast and Robust Models for Biomedical Natural Language Processing. In *Proceedings of the 18th BioNLP*

*Workshop and Shared Task*, pages 319–327, Florence, Italy. Association for Computational Linguistics.

Rotmensch, M., Halpern, Y., Tlimat, A., Horng, S., and Sontag, D. (2017). Learning a health knowledge graph from electronic medical records. *Scientific Reports*, 7(1):5994.

SNOMED International (2024). SNOMED CT. Accessed: 2024-10-28.

Wu, X., Zeng, Y., Das, A., Jo, S., Zhang, T., Patel, P., Zhang, J., Gao, S.-J., Pratt, D., Chiu, Y.-C., et al. (2024). regulogpt: Harnessing gpt for knowledge graph construction of molecular regulatory pathways. *bioRxiv*.

Yu, S., Yuan, Z., Xia, J., Luo, S., Ying, H., Zeng, S., Ren, J., Yuan, H., Zhao, Z., Lin, Y., et al. (2022). Bios: An algorithmically generated biomedical knowledge graph. *arXiv preprint arXiv:2203.09975*.

Zhang, R., Hristovski, D., Schutte, D., Kastrin, A., Fiszman, M., and Kilicoglu, H. (2021). Drug repurposing for covid-19 via knowledge graph completion. *Journal of Biomedical Informatics*, 115:103696.

Zhang, Y., Li, Y., Cui, L., Cai, D., Liu, L., Fu, T., Huang, X., Zhao, E., Zhang, Y., Chen, Y., et al. (2023). Siren's song in the ai ocean: a survey on hallucination in large language models. *arXiv preprint arXiv:2309.01219*.
